# Surveillance of COVID-19 in a Vaccinated Population: A Rapid Literature Review

**DOI:** 10.1101/2021.11.05.21265763

**Authors:** Oluwaseun Egunsola, Brenlea Farkas, Jordyn Flanagan, Charleen Salmon, Liza Mastikhina, Fiona Clement, on behalf of the University of Calgary Health Technology Assessment Unit

## Abstract

**Objectives:** With the availability of COVID-19 vaccines, public health focus is shifting to post-vaccination surveillance to identify breakthrough infections in vaccinated populations. Therefore, the objectives of these reviews are to identify scientific evidence and international guidance on surveillance and testing approaches to monitor the presence of the virus in a vaccinated population.

**Method:** We searched Ovid MEDLINE®, including Epub Ahead of Print, In-Process & Other Non-Indexed Citations, Embase, EBM Reviews - Cochrane Central Register of Controlled Trials, and EBM Reviews - Cochrane Database of Systematic Reviews. We also searched the Web of Science Core Collection. A grey literature search was also conducted. This search was limited to studies conducted since December 2020 and current to June 13th, 2021. There were no language limitations. COVID-19 surveillance studies that were published after December 2020 but did not specify whether they tested a vaccinated population were also considered for inclusion.

For the international guidance review, a grey literature search was conducted, including a thorough search of Google, websites of international government organizations (e.g., Center for Disease Control and Prevention [CDC], World Health Organization [WHO]), and McMaster Health Forum (CoVID-END). This search was primarily examining surveillance guidance published since December 2020 (to capture guidance specific to vaccinations) and any relevant pre-December 2020 guidance.

**Results:** Thirty-three studies were included for data synthesis of scientific evidence on surveillance of COVID-19. All the studies were published between April and June 2021. Twenty-one studies were from peer-reviewed journals. Five approaches to monitoring post-vaccination COVID-19 cases and emerging variants of concern were identified, including screening with reverse transcriptase polymerase chain reaction (RT-PCR) and/or a rapid antigen test, genomic surveillance, wastewater surveillance, metagenomics, and testing of air filters on public buses. For population surveillance, the following considerations and limitations were observed: variability in person-to-person testing frequency; lower sensitivity of antigen tests; timing of infections relative to PCR testing can result in missed infections; large studies may fail to identify local variations; and loss of interest in testing by participants in long follow-up studies.

Through comprehensive grey literature searching, 68 international guidance documents were captured for full-text review. A total of 26 documents met the inclusion criteria and were included in our synthesis. Seven overarching surveillance methods emerged in the literature. PCR-testing was the most recommended surveillance method, followed by genomic screening, serosurveillance, wastewater surveillance, antigen testing, health record screening, and syndromic surveillance.

**Conclusion:** Evidence for post-vaccination COVID-19 surveillance was derived from studies in partially or fully vaccinated populations. Population PCR screening, supplemented by rapid antigen tests, was the most frequently used surveillance method and also the most commonly recommended across jurisdictions. Most recent guidance on COVID-19 surveillance is not specific to vaccinated individuals, or it is in effect but has not yet been updated to reflect that. Therefore, more evidence-informed guidance on testing and surveillance approaches in a vaccinated population that incorporates all testing modalities is required.

**EXECUTIVE SUMMARY:** *Objectives:* With the availability of COVID-19 vaccines, public health focus is shifting to post-vaccination surveillance to identify breakthrough infections in vaccinated populations. Therefore, the objectives of these reviews are to: 1) identify scientific evidence on surveillance and testing approaches to monitor the presence of the virus in a vaccinated population and determine how these influence testing strategies; 2) identify international guidance on testing and surveillance for COVID-19 and its variants of concern in a vaccinated population; and 3) identify emerging technologies for surveillance.

*Design:* A rapid review was conducted to identify scientific evidence on COVID-19 surveillance and testing approaches, and a targeted literature review was conducted on international guidance.

*Method:* We searched Ovid MEDLINE®, including Epub Ahead of Print, In-Process & Other Non-Indexed Citations, Embase, EBM Reviews - Cochrane Central Register of Controlled Trials, and EBM Reviews - Cochrane Database of Systematic Reviews. We also searched the Web of Science Core Collection. We performed all searches on June 13, 2021. A grey literature search was also conducted, including: MedRxiv, Google, McMaster Health Forum (COVID-END), and websites of international government organizations (e.g., Center for Disease Control and Prevention [CDC], World Health Organization [WHO]). This search was limited to studies conducted since December 2020 and current to June 13th, 2021. There were no language limitations. COVID-19 surveillance studies that were published after December 2020 but did not specify whether they tested a vaccinated population were also considered for inclusion. For the international guidance review, a grey literature search was conducted, including a thorough search of Google, websites of international government organizations (e.g., Center for Disease Control and Prevention [CDC], World Health Organization [WHO]), and McMaster Health Forum (CoVID-END). This search was primarily examining surveillance guidance published since December 2020 (to capture guidance specific to vaccinations) and any relevant pre-December 2020 guidance. Although the primary focus was on surveillance guidance in a vaccinated population, guidance that was published after December 2020 but was not vaccine-specific was also considered for inclusion; it was assumed that this guidance was still in effect and was not yet updated. There were no language limitations. A patient partner was engaged during the co-production of a plain language summary for both the rapid review of primary literature and the review of international guidance.

*Results:* Thirty-three studies were included for data synthesis of scientific evidence on surveillance of COVID-19. All the studies were published between April and June 2021. Twenty-one studies were from peer-reviewed journals. Five approaches to monitoring post-vaccination COVID-19 cases and emerging variants of concern were identified including, screening with reverse transcriptase polymerase chain reaction (RT-PCR) and/or a rapid antigen test, genomic surveillance, wastewater surveillance, metagenomics, and testing of air filters on public buses. Population surveillance with RT-PCR testing and/or rapid antigen testing was utilized in 22 studies, mostly in healthcare settings, but also in long-term care facilities (LTCFs) and in the community. The frequency of testing varied depending on whether there was an outbreak. For population surveillance, the following considerations and limitations were observed: studies with discretionary access to testing have highly variable person-to-person testing frequency; antigen tests have lower sensitivity, therefore some positive cases may be missed; timing of infections relative to PCR testing as well as the sensitivity of the tests can result in missed infections; large sample sizes from multicentre studies increase generalizability, but fail to identify local variations from individual centres; with electronic database surveillance, it is difficult to confirm whether patients with a breakthrough infection and a previous positive SARS-CoV-2 test result had a true reinfection or had prolonged shedding from the previous infection; and participants lose interest in studies with long follow-up, with decrease in testing rates over time. Six wastewater surveillance and three genomic surveillance studies were identified in this review. A number of benefits such as, good correlation with clinical data, ability to predict major outbreaks, and rapid turnaround time were observed with wastewater surveillance. However, challenges such as, inconsistencies in variant representation depending on where the samples were taken within the community, differences in the capacity of wastewater to predict case numbers based on the size of the wastewater treatment plants, and cost, were noted. Emerging technologies like viral detection in public transport filters, novel sampling, and assay platforms were also identified. Through comprehensive grey literature searching, 68 international guidance documents were captured for full-text review. A total of 26 documents met the inclusion criteria and were included in our synthesis. Most were not specific to vaccinated populations but reported on a surveillance method of COVID-19 and were therefore included in the review; it was assumed that they were still in effect but have not yet been updated. Eleven countries/regions were represented, including Australia, Brazil, France, Germany, India, New Zealand, Spain, United Kingdom, United States, Europe, and International. All of the guidance documents included surveillance methods appropriate for community settings. Other settings of interest were healthcare settings, including hospitals and primary care centres, long-term care facilities, points of entry for travel, schools, and other sentinel sites (e.g., prisons and closed settings). Seven overarching surveillance methods emerged in the literature. PCR-testing was the most recommended surveillance method, followed by genomic screening, serosurveillance, wastewater surveillance, antigen testing, health record screening, and syndromic surveillance. Only one document (published by Public Health England) was identified that provided guidance on surveillance specific to vaccinated populations. The document outlined a plan to surveil and monitor COVID-19 in vaccinated populations through a series of targeted longitudinal studies, routine surveillance, enhanced surveillance, use of electronic health records, surveillance of vaccine failure (including follow-up with viral whole genome sequencing), and sero-surveillance (including blood donor samples, routine blood tests, and residual sera).

*Conclusion:* Evidence for post-vaccination COVID-19 surveillance was derived from studies in partially or fully vaccinated populations. Population PCR screening, supplemented by rapid antigen tests, was the most frequently used surveillance method and also the most commonly recommended across jurisdictions. The selection of testing method and the frequency of testing was determined by the intensity of the disease and the scale of testing. Other common testing methods included wastewater surveillance and genomic surveillance. A few novel technologies are emerging, however, many of these are yet to be utilized in the real-world setting. There is limited evidence-based guidance on surveillance in a vaccinated population. Most recent guidance on COVID-19 surveillance is not specific to vaccinated individuals, or it is in effect but has not yet been updated to reflect that. Therefore, more evidence-informed guidance on testing and surveillance approaches in a vaccinated population that incorporates all testing modalities is required.

*Protocol/Topic Registration:* PROSPERO-CRD42021261215.

**Key Definitions:** **Antigen:** a foreign protein which induces an immune response in the body, especially the production of antibodies

**Fully vaccinated**: refers to individuals who have received complete dosage of a given vaccine

**Partially vaccinated:** refers to individuals who have received an incomplete dosage of a given vaccine

**Sero-surveillance:** estimation of antibody levels against infectious diseases

**Surveillance:** ongoing systematic collection, analysis, and interpretation of health data that are essential to the planning, implementation, and evaluation of public health practice

**Variants of Concern:** a variant for which there is evidence of an increase in transmissibility and/or more severe disease

**Variants:** virus with a permanent change in its genetic sequence

## Introduction

Coronavirus disease (COVID-19) is caused by severe acute respiratory syndrome coronavirus 2 (SARS-CoV-2). As of June 2021, there have been more than 179,000,000 confirmed cases of COVID-19, which have resulted in more than 3,800,000 confirmed deaths worldwide.^1^ Numerous randomized controlled trials (RCTs) and real-world observational studies have found vaccines to be safe and effective at preventing COVID-19.^2^ At the time of writing, more than 2,700,000 vaccine doses have been administered across the world, with several countries (e.g., Israel, the UK) approaching the 70% benchmark of having their population fully vaccinated with the goal of reaching herd immunity.^1, 3^

As the number of partially and fully vaccinated people continues to grow, countries may be pivoting their population-level surveillance methods to capture the presence and resurgence of COVID-19 and its variants of concern (VOCs). RT-PCR testing of nose and throat swabs is a widely used method to identify new cases of COVID-19; however, the ability of RT-PCR testing to slow viral spread may be impacted by slow laboratory turnaround times and restricted availability of the tests.^4^ As a result, there may be interest in alternative population-level testing modalities that can detect the presence and resurgence of the virus in a setting before an outbreak.

This rapid review aims to answer the following research questions:

1. What scientific evidence exists on surveillance approaches to monitor the presence of the virus in a fully vaccinated population (i.e., monitoring for resurgence and variants of concern through wastewater surveillance and metagenomics, population screening with rapid antigen testing)? How does this influence testing strategies?

a. What technologies are emerging to identify infection caused by variants of concern in a vaccinated population?
2. What international guidance exists on testing and surveillance for SARS-CoV-2 and its variants of concern in a fully vaccinated population?

Recognizing that the evidence on surveillance in fully vaccinated populations may be limited, this review also includes literature assessing surveillance broadly in a partially vaccinated population to ensure that all relevant literature is captured.

## Section 1: Scientific Evidence on Surveillance Methods in a Vaccinated Population

### Methods

This rapid review is registered in the International Prospective Register of Systematic Reviews (PROSPERO), number CRD42021261215. An experienced medical information specialist developed and tested the search strategies through an iterative process in consultation with the review team. Another senior information specialist peer reviewed the MEDLINE strategy prior to execution using the PRESS Checklist.^5^ Using the multifile option and deduplication tool available on the OVID platform, we searched Ovid MEDLINE®, including Epub Ahead of Print, In-Process & Other Non-Indexed Citations, Embase, EBM Reviews - Cochrane Central Register of Controlled Trials, and EBM Reviews - Cochrane Database of Systematic Reviews. We also searched the Web of Science Core Collection. We performed all searches on June 13, 2021.

The strategies utilized a combination of controlled vocabulary (e.g., “COVID-19”, “Epidemiological Monitoring”, “Population Surveillance”) and keywords (e.g., “nCoV”, “vaccinated”, “surveillance”). Vocabulary and syntax were adjusted across the databases (full search strategies included in Appendix A). No language restrictions were applied but results were limited to the publication years 2020 to the present. Results were downloaded and deduplicated using EndNote version 9.3.3 (Clarivate Analytics) and uploaded to Microsoft Word.

A grey literature search was also conducted, including: MedRxiv, Google, McMaster Health Forum (CoVID-END), and websites of international government organizations (e.g., Center for Disease Control and Prevention [CDC], World Health Organization [WHO]). This search was limited to studies conducted since December 2020 and current to June 13^th^, 2021. There were no language limitations.

A screening form based on the eligibility criteria was prepared. Citations identified as potentially relevant from the literature search were screened by single reviewer across a team of four reviewers and subsequently read in full text by two reviewers and assessed for eligibility based on the criteria outlined below (Table 1). Discrepancies were resolved by discussion or by a third reviewer. Reference lists of included studies were hand searched to ensure all relevant literature is captured.

**Table 1.**
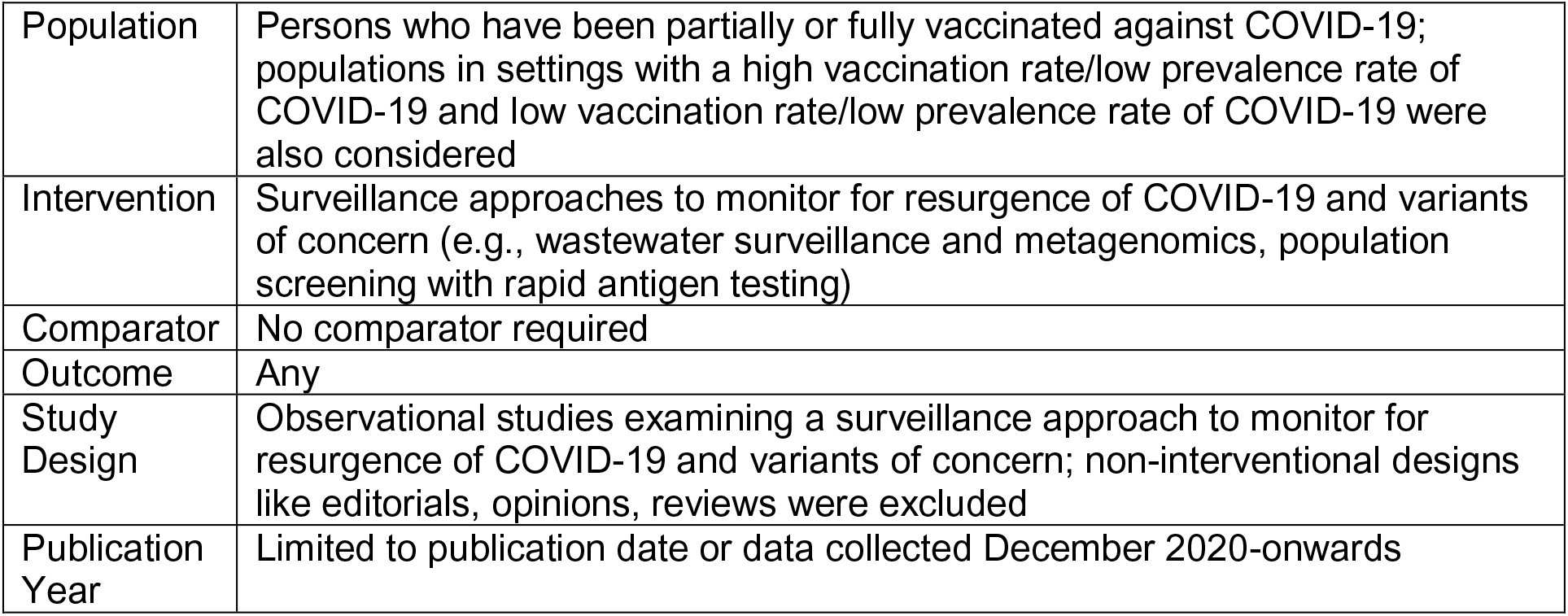
Criteria for Inclusion of Scientific Evidence on Surveillance

A standardized data extraction sheet was used to extract the month and year of publication, country, study design, surveillance method, variant surveillance, dates of enrollment, vaccination status, population, setting, primary outcomes, and implementation considerations. All reviewers completed a calibration exercise whereby data from two sample studies were extracted by all four reviewers and areas of disagreement were discussed. Data were extracted by one reviewer.

Given the rapid nature of this request, a formal risk of bias assessment was not conducted. A high-level discussion of quality of the evidence (i.e., peer-reviewed vs. preprints) is included below.

Due to the heterogeneity in study designs and outcomes across included studies, data were synthesized narratively; a meta-analysis was not conducted. A high-level summary of the different surveillance methods, populations assessed by the methods, and outcomes is presented in the next section. A patient partner was engaged during the co-production of a plain language summary, which is presented in a separate document.

### Results

A total of 1197 articles were identified from database search. After removing duplicates, 914 unique citations were included; 90 of which were assessed in full text articles. Thirty-three studies were included for data synthesis (Figure 1). All the studies were published between April and June 2021. Thirteen were national studies; there were seven regional and city-wide studies each; five were single-centre studies (e.g., hospitals and long-term care facilities [LTCFs]); and one was an international study. Sixteen studies were conducted in the USA, four were from England, three each from Israel and Spain, two from Italy, and one study from each of the following: the UK, Canada, India, Indonesia, and Uruguay. The majority of the studies (n=21) were from peer-reviewed journals (Table 2).

**Figure 1:**
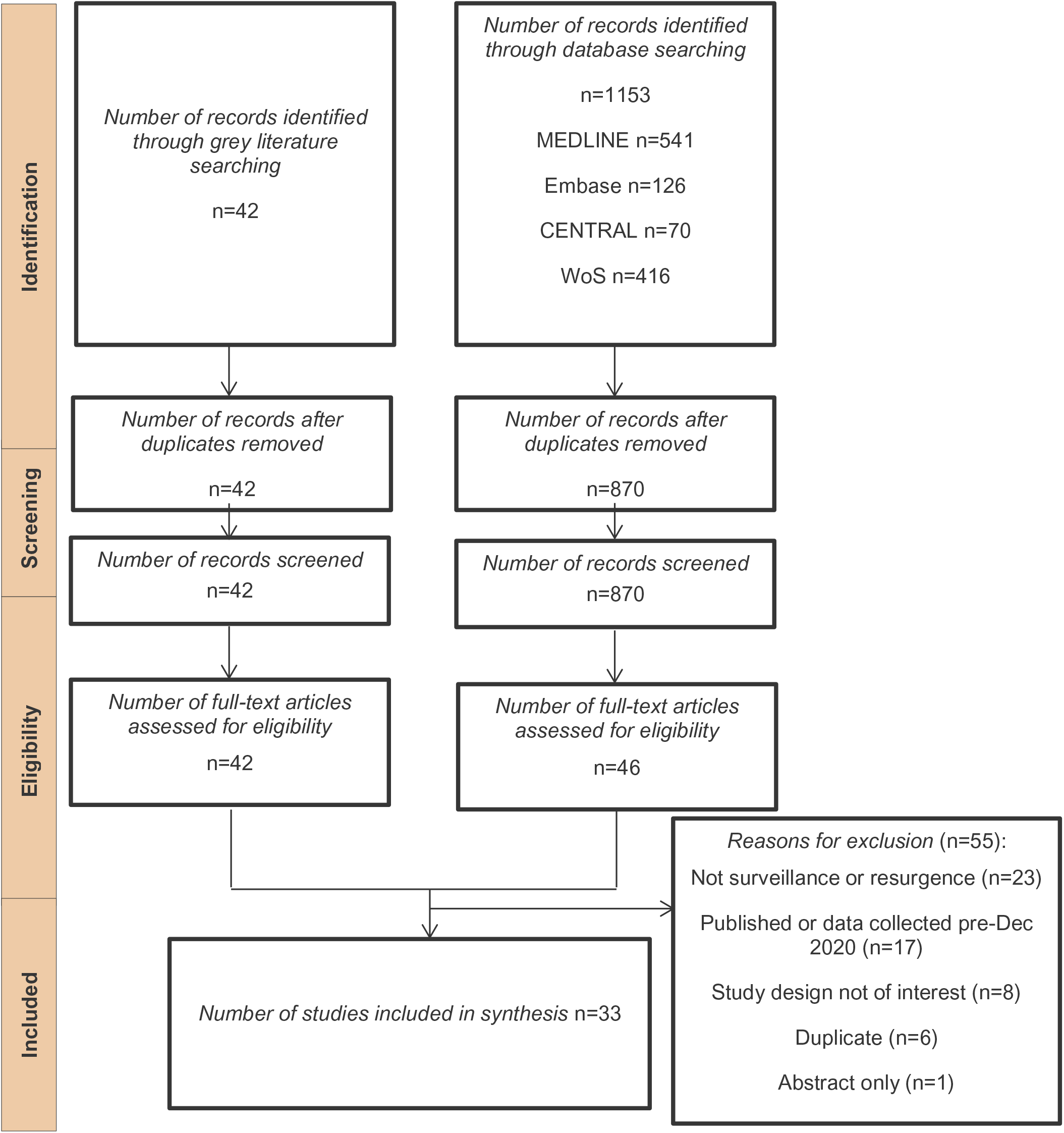
Flowchart of Studies Included in the Review of Scientific Evidence

**Table 2:** Surveillance Studies Involving Vaccinated Populations

| Author (Country) | Setting/Scope | Surveillance method | Highlights from Surveillance |
| --- | --- | --- | --- |
| **Population RT-PCR or Rapid Antigen Screening** |  |  |  |
| Angel et al.<sup>12</sup><br>(Israel)<br>Peer - Reviewed | Healthcare/Single Hospital | Routine screening of HCWs with potential exposure using nasopharyngeal swabs and RT-PCR based virus detection | This was a retrospective cohort study involving fully vaccinated populations. The frequency of testing varied from person to person and health workers were able to access testing at their discretion. No variant surveillance was conducted. |
| Baj et al.<sup>11</sup><br>(Italy)<br>Peer-Reviewed | Healthcare/Single Hospital | Nasopharyngeal swabs of HCWs with RT-PCR testing. Testing frequency not reported. | This was a passive surveillance study describing a series of 11 voluntarily reported cases in fully vaccinated individuals. Spike gene sequencing showed they tested positive for different strains with 7 of 11 testing for B.1.1.7 (x7 patients) variant. |
| Bouton et al.<sup>10</sup><br>(USA)<br>Preprint | Healthcare/Single Hospital | Asymptomatic RT-PCR testing was available to HCWs for workplace exposures, following out-of-state travel, and per request. 68 single nucleotide variants identified post at least one dose vaccine | This was a prospective cohort study of partial and fully vaccinated individuals showing a positive impact of COVID-19 vaccines on SARS-CoV-2 case rates was seen. Postvaccination isolates did not show unusual genetic diversity or selection for mutations of concern. Variant surveillance found no early evidence of specific spike mutations or mutations associated with neutralizing vaccine escape. |
| Cucunawangsi h et al.<sup>9</sup><br>(Indonesia)<br>Grey Literature (Report) | Healthcare/Single Hospital | Symptomatic and post-exposure RT-PCR and antigen testing of HCWs | This was a retrospective study involving fully vaccinated individuals. Through passive and active surveillance, breakthrough cases were identified in 13 of the 1040 fully vaccinated HCWs. Because of the reduced sensitivity of antigen testing in asymptomatic populations, it was possible that some asymptomatic cases were not identified. |
| Hall et al.<sup>7</sup><br>(England) | Healthcare/National | Twice weekly rapid antigen testing of HCWs per government guidelines. RT- | This was a prospective cohort study in partial or fully vaccinated individuals. It was possible that infections |
| Peer-Reviewed |  | PCR testing of anterior nasal swabs or combined nose and oropharyngeal swabs every 2 weeks and monthly antibody testing. | were missed during follow-up, depending on timing of infection relative to PCR testing and PCR sensitivity. |
| Jacobson et al.<sup>6</sup> (USA)<br>Preprint | Healthcare/National | Self-monitoring for symptoms. If symptoms were present, RT-PCR testing was conducted. Some HCWs participated in voluntary weekly asymptomatic RT-PCR testing. | This is a retrospective cohort study of partially vaccinated individuals. The great majority of post-vaccine SARS-CoV-2 occurred prior to the expected onset of full, vaccine-derived immunity. |
| Jones et al.<sup>15</sup> (England)<br>Peer-Reviewed | Healthcare/Single Hospital | HCWs or HCW household member could self-refer for RT-PCR testing if symptoms develop. HCWs had weekly asymptomatic testing at temporary on-site 'Pods' and via oropharynx and anterior nasal cavity self-swabbing. | This was a retrospective cohort study of partially vaccinated individuals. Investigating asymptomatic infections means that the date of infection could be earlier than the test date for the virus. |
| Sansone et al.<sup>8</sup> (Italy)<br>Peer Reviewed | Healthcare/Regional | RT-PCR testing on rhino-pharyngeal swabs, followed by gene sequencing in positive vaccinated HCWs. Testing frequency not reported. | This was a retrospective cohort study conducted when 78% of the workforce had been fully vaccinated, while a recrudescence wave of the SARS-CoV-2 pandemic hit Lombardy and particularly the Brescia County, where the B.1.1.7 variant was highly prevalent. |
| Tyagi et al.<sup>16</sup> (India)<br>Peer-Reviewed | Healthcare/Single Hospital | RT-PCR and rapid antigen testing. Testing frequency not reported. | A passive surveillance approach was utilized in order to detect breakthrough cases following 94% full vaccination of work-force. There was no report on variant surveillance |
| Tang et al.<sup>13</sup> (USA)<br>Peer-Reviewed | Healthcare/Single Hospital | Asymptomatic RT-PCR testing of mid-turbinate samples of hospital employees performed at least weekly | This was a prospective cohort study in partially or fully vaccinated individuals. The follow-up time was short. |
| Lutrick et al.<sup>14</sup> (USA) | Healthcare and Essential workers/Regional | Weekly RT-PCR testing of mid-turbinate samples. If participant experiences self- | Participants were prompted to provide samples at regular intervals via SMS. The study is ongoing with |
| Preprint |  | reported symptoms, asked to provide additional nasal swab specimen. Each week, all participants were contacted via text message on their predesignated surveillance day. At the end of each text message exchange, the participant was reminded to collect a weekly specimen on their assigned day for collection. | no findings yet. The population of fully vaccinated Arizonans at the time of publication was not reported. Outcomes and generalisability could likely be biased due to healthy worker effect. Information from participants is self-reported or self-collected, therefore possible recall bias may be present. |
| Thompson et al<sup>41</sup><br>(USA)<br>Peer-Reviewed | Healthcare and Essential workers/National | Self-collected a mid-turbinate nasal swab weekly, regardless of COVID-19–associated illness symptom status. Collected an additional nasal swab and saliva specimen at the onset of COVID-19–associated illness for RT-PCR and antigen testing | An active symptom surveillance preceded nasal swabbing. Study was a longitudinal cohort conducted in eight cities across the USA. The cities were expected to have varying vaccination rates (not reported). |
| Rudolph et al<sup>20</sup><br>(USA)<br>Peer-Reviewed | Long Term Care/National | RT-PCR and rapid antigen testing. Testing frequency not reported, but positive cases were summarized weekly. | This was a retrospective cohort analysis of 130 department of veteran affairs community living centres (CLCs). The community nursing home data are self-reported, but do not report on the testing and vaccination practices of nursing homes. There are also major differences in patient populations between VA CLCs and CMS-certified nursing homes. Staff vaccination data was lacking. The large geographic spread of the VA system allowed a nationwide sampling but limited the detail on local variation in practices. |
| De Salazar et al<sup>21</sup><br>(Spain)<br>Preprint | Long Term Care/Regional | All contacts among staff and residents were screened using molecular test (PCR or antigen test) immediately upon confirmation of an index infection in a facility; further, all staff and residents were regularly screened independently of whether the individuals show symptoms or not; public health guidelines required to screen staff every 2-4 weeks depending on the population size where the LTCFs is located | This was a retrospective study of fully vaccinated individuals. Analysis was conducted at county level rather than facility-level. |
| White et al.<sup>18</sup><br>(USA)<br>Peer-Reviewed | Long Term Care/National | Residents tested with RT-PCR or antigen every 3-7 days when there was a confirmed cases in the facility, and tested if they were symptomatic or potential exposure | Post partial/full vaccination cases were monitored using electronic health record data from Genesis Health- Care, a large long-term care provider in the United States (280 nursing homes in 21 States). |
| Domi et al.<sup>17</sup><br>(USA)<br>Peer-Reviewed | Long Term Care/National | In accordance with CMS (Centers for Medicare and Medicaid services) regulations, nursing homes in the same county were required to test all their staff and residents at the same frequency based on the county COVID-19 rate | In this retrospective study, cases were assessed approximately two months into vaccination. The level of vaccination at these facilities was unclear. CMS required all nursing homes to test residents and staff at the same frequency dictated by the rate of COVID-19 in the community not in the facility. |
| Shrotri et al.<sup>19</sup><br>(England)<br>Preprint | Long Term Care/National | LTCF residents underwent monthly routine RT-PCR testing, and if an LTCF outbreak was suspected, local public health teams organised PCR testing for all residents upon notification and 7 days later. Individuals who tested positive were not re-tested for the following 90 days unless they developed new COVID-19 symptoms | This prospective study analyzed cases of COVID-19 during the first three months of vaccine roll-out in the UK. A benefit of this study was access to high-quality routine data for a large, well-defined cohort of LTCF residents who were tested regularly for SARS-CoV-2 throughout follow-up. Also, cohort included a range of LTCF types making findings to be generalisable across LTCF resident population. |
| Cavanaugh et al.<sup>22</sup><br>(USA)<br>Peer-Reviewed | Long Term Care/Single facility | Twice-weekly RT-PCR testing of all HCP. Once the outbreak was confirmed, daily rapid point-of-care antigen testing of all residents, regardless of symptoms, was added to the twice-weekly PCR | This was an outbreak investigation report, which described the pre-outbreak surveillance practices and an enhanced active surveillance after the outbreak. 90% and 53% of staff and residents had been fully vaccinated before the outbreak. During |
|  |  | testing. Additional specimens were collected the same day for RT-PCR confirmation of positive antigen test results. One week after the outbreak was identified, resident antigen testing was reduced to three times weekly, then to twice weekly after no additional cases were identified for 1 week. | the outbreak, 46 COVID-19 cases were identified, including cases in 26 residents (18 fully vaccinated) and 20 HCP (four vaccinated). |
| Teran et al<sup>23</sup><br>(USA)<br>Peer-Reviewed | Long Term Care/City | Monitor infections using a data triangulation method that matches the SARS-CoV-2 test results from RT-PCR and antigen tests reported to the Illinois' National Electronic Disease Surveillance System with facility-reported line lists of SARS-CoV-2 test results from routine screening testing. | The Chicago Department of Public Health (CDPH) began matching records to Illinois' Comprehensive Automated Immunization Registry Exchange to identify breakthrough infections. After identifying SARS-CoV-2 infection in a resident 16 days after receipt of a second vaccine dose, CDPH initiated an investigation to quantify breakthrough infections across all facilities. Confirming whether patients with a breakthrough infection and a previous positive SARS-CoV-2 test result had a true reinfection or represented persons with prolonged shedding from previous infection was not possible. Breakthrough infections might be underestimated because of data entry errors or delayed surveillance reporting. |
| Haas et al<sup>25</sup><br>(Israel)<br>Peer-Reviewed | Community/National | Voluntary RT-PCR testing based on travel history, symptoms and hospitalization. | A prospective cohort design was implemented using the Israeli national surveillance data from the first 4 months of the nationwide vaccination campaign. Only data from fully vaccinated was analyzed. Daily reporting from laboratories to the national database was legally required. |
| Riley et al. <sup>24</sup><br>(England)<br>Preprint | Community/National | Collected a self-administered throat and nose swab sample and questionnaire data from a random sample of the population in England at ages 5 years | This was the 11<sup>th</sup> iteration of a surveillance study which was conducted about 5 months into England's vaccination program (April 15 to May 3, 2021). Prevalence of new cases were compared with the |
|  |  | and above. Used the National Health Service (NHS) register of patients to select the sample aiming to obtain similar numbers of participants in each of the 315 lower tier local authorities (LTLAs) in England.B.1.1.7, B.1.617.2 | preceding iteration. The authors observed a decline in response rate compared with preceding rounds and there were concerns regarding the representativeness of the samples collected. Based on sequence data for positive samples for which a lineage could be identified, estimated that that 92.3% of infections were from the B.1.1.7 lineage and 7.7% from the B.1.617.2 lineage |
| Munitz et al.<sup>26</sup><br>(Israel)<br>Peer-Reviewed | Community and Long-Term Care/National | Approximately 300,000 individual tests collected from Israeli nursing houses and from random "Drive and Check" SARSCoV-2 test complexes. | Monitored the dynamics of the spread of the B.1.1.7 SARS-CoV-2 variant in Israel, using samples collected two months into the vaccine roll-out. The study demonstrated that the B.1.1.7 variant was 45% more transmissible than the wild-type strain in Israel. |
| **Genomic Surveillance** |  |  |  |
| Thompson et al.<sup>27</sup><br>(USA)<br>Peer-Reviewed | Community/City | All RT-PCR positive SARS-CoV-2 specimens, from 9 COVID express laboratories serving New York City, with a cycle threshold (Ct) value <32 underwent whole genome sequencing. | Sequencing was done for samples collected during the first four months of vaccination. The number of persons with reinfection or breakthrough infection whose specimens underwent whole genome sequencing was low, limiting the statistical power to detect modest increases in immune escape that could have a substantial impact on public health. Improved capacity for genomic surveillance, establishment of automated and efficient exchange of WGS data, and integration with population-based clinical and epidemiologic data would enable the rapid characterization of emerging SARS-CoV-2 variants. |
| Rego et al.<sup>28</sup><br>(Uruguay) | Community/National | Daily sampling of four diagnostic laboratories was included in the network, that were able to process all | Showed that VOC P.1 was introduced in Uruguay at multiple times over a period of increasing mobility. |
| Preprint |  | together more than 3,000 nasopharyngeal samples per day. Of those, between 200 – 300 SARS-CoV2 positive samples were received weekly for qPCR-VOC analysis and 50% of them were further processed for SARS-CoV-2 genome sequencing. Positive RNA samples were obtained from hospitalised cases and outpatient cases collected from all over the country. P1 variants | Samples were collected within the first 3 months of vaccine availability. |
| Wang et al.<sup>29</sup> (USA)<br>Peer-Reviewed | Community/City | Upper respiratory swab specimens collected from patients as part of routine clinical care. Samples testing positive for SARS-CoV-2 with RT-qPCR Ct ≤ 30 or transcription-mediated amplification relative light units (RLU) ≥ 1,100 during this period were subject to multiplex allele-specific genotyping. | Specimen were collected during the first three months of vaccine availability in the USA. The assay revealed rapid emergence of L452R in population, with a prevalence of 24.8% in December 2020 that increased to 62.5% in March 2021. This approach can be adapted for emerging mutations and implemented in laboratories already conducting SARS-CoV-2 NAAT using existing resources and extracted nucleic acid |
| **Wastewater Surveillance** |  |  |  |
| Peterson et al.<sup>34</sup> (Canada)<br>Preprint | Community/Regional | A 24-hr composite wastewater sample was collected three times per week. Wastewater was collected from fifteen urban (Vancouver, Edmonton, Toronto, Montreal and Halifax) wastewater treatment plants (WWTP). Wastewater was also collected from 7 lift stations from remote communities in the Northwest Territories that are not part of the CWS. | Variants of concern were not detected in the Territorial communities, suggesting the absence of variants of concern SARS-CoV-2 cases in those communities. Percentage of variant remained low throughout the study period in the majority of the sites tested. However, the Toronto sites showed a marked increase from ~25% to ~75% over the study period. |
| Sanjuan et al.<sup>31</sup><br>(Spain)<br>Preprint | Community/City | Wastewaters from the Pinedo and Quart-Benàger Wastewater Treatment plant in the metropolitan area of Valencia (Spain) were sampled (200 mL) by grabbing or taking 4 h composite samples between 7 and 11 am. Sampling was carried out on 129 different days spanning from April 16, 2020, to March 9, 2021. | Time series were similar for wastewaters data and declared cases, but wastewater RNA concentrations exhibited transient peaks that were not observed in declared cases and preceded major outbreaks by several weeks. B.1.1.7 identified. |
| Fitzgerald et al.<sup>35</sup><br>(UK)<br>Peer-Reviewed | Community/National | Sites were sampled once a week for RT-PCR testing | There were significant differences between wastewater treatment plants in their capacity to predict case numbers based on influent viral RNA load, with the limit of detection ranging from twenty-five cases for larger plants to a single case in smaller plants. |
| Fuqua et al.<sup>33</sup><br>(USA)<br>Preprint | Community/Regional | RNA isolated from wastewater samples was used to quantify SARS-CoV-2 and analyze the genetic variation through high-throughput sequencing. Bioinformatics approaches were used to rapidly identify single nucleotide genetic alterations, which were compared with known variants of interest and concern. | Differences in the scale of sample pooling in the community revealed unanticipated inconsistencies in variant representation. Variants observed in smaller catchment areas, such as neighbourhood manhole locations, were not observed in downstream treatment plants, suggesting catchment size or population could impact the ability to detect diversity. |
| Kantor et al.<sup>32</sup><br>(USA)<br>Preprint | Community/City | RT-PCR testing once a week | The cost for laboratory buildout was approximately \$100,000 USD in capital equipment and took 3 months to complete. The laboratory used the kit-free sewage, salt, silica, and SARS-CoV-2 (4S) direct RNA extraction method and reverse transcription quantitative polymerase chain reaction (RT-qPCR) with a per-sample cost (consumables and reagents) of around \$25 USD. |
|  |  |  | Turnaround time (number of days between sample collection and results was 1.6 days |
| Carcereny et al.<sup>30</sup><br>(Spain)<br>Peer-Reviewed | Community/National | Weekly RT-PCR assay and S gene sequencing with Single nucleotide polymorphism (SNP) identification. | Wastewater-based tracking showed good correlation with clinical data providing information at local level. Study highlighted applicability of Rt-PCR based strategy to track specific mutations of VOC as soon as they are identified by clinical sequencing. The relative proportion of B.1.1.7 variant in wastewater could be estimated for 91% of positive samples |
| **Testing air filters on public buses** |  |  |  |
| Hoffman et al.<sup>36</sup><br>(USA)<br>Preprint | Community/Regional | Individual buses were selected to be sampled via a convenience sampling approach based on which buses could be made available at the depot on a regular basis between 7:00-9:00 AM for sample retrieval. Retrieved filters in the existing HVAC system were sampled. Sample extraction for RT-PCR testing was performed within the same day of the sample collection from metro buses. | SARS-CoV-2 RNA was detected in 14% of public bus filters tested. There was an overlap between pre-and post-vaccination testing periods in this study. Mask mandates were in effect for riders during the sample period, likely reducing the number of viral particles from infected riders landing on the filters. |
| **Metagenomics** |  |  |  |
| Quinonez et al.<sup>37</sup><br>Peer-Reviewed | NextStrain.org/International | Computational modelling of genomic sequences. | Predicted that the rate of B.1.1.7 lineage is going to sharply decrease (near to zero) in coming months. In contrast, the frequency of other variants, B.1.351, B.1.617, and P.1 will gradually increase. The authors suggested a cautious generalizability because of the lack of validation in an experimental study. |

Abbreviations: HCW: Healthcare Worker, VOC: Variant of Concern, RT-PCR: Reverse Transcriptase Polymerase Chain Reaction

#### Surveillance Methods for COVID-19 Cases

Five approaches to monitoring post-vaccination COVID-19 cases and emerging variants of concern were identified in this review. These include population screening with reverse transcriptase polymerase chain reaction (RT-PCR) and or a rapid antigen test, genomic surveillance, wastewater surveillance, metagenomics, and testing of air filters on public buses (Figure *2*).

**Figure 2:**
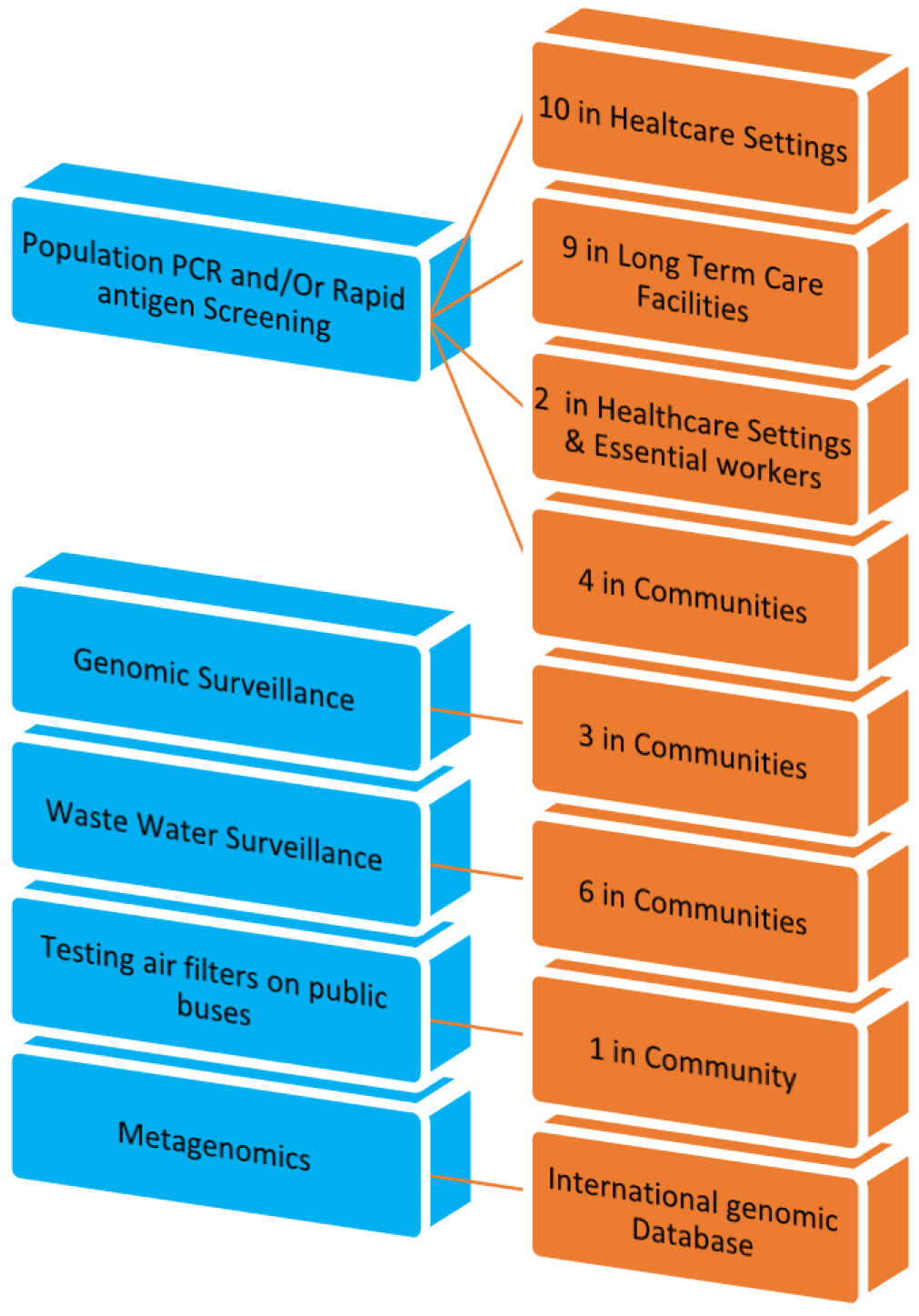
Summary of Scientific Evidence on Surveillance Methods

##### Population Screening with RT-PCR and or Rapid Antigen Testing

RT-PCR testing and or rapid antigen testing was utilized in 22 studies. The majority of these studies were in healthcare settings (10 studies), seven were in LTCFs, and two in the community. In two studies, surveillance was conducted among healthcare and essential workers, while another two studies involved LTCFs and the community (Table 2).

###### Healthcare Setting

The studies involving healthcare workers (HCWs) were mostly conducted at single hospitals (7 studies); while two studies were national,^6, 7^ and one was a regional study.^8^ In some studies, routine post-exposure screening of symptomatic or asymptomatic HCWs using RT-PCR based testing of samples obtained from nasal or oropharyngeal swabs were implemented;^9–12^ while some other studies implemented periodic (typically weekly) asymptomatic screenings.^6, 13–15^ Although the majority of the surveillance involving HCWs utilized only RT-PCR testing, two studies used a combination of RT-PCR and rapid antigen testing. One was a biweekly national surveillance study of HCWs in England,^7^ and the other was at a single hospital in India.^16^ Seven of the hospital-based studies also implemented genetic sequencing to identify variants following a positive RT-PCR test. All the studies identified breakthrough cases among vaccinated individuals, some sequenced for VOCs, while also demonstrating the effectiveness of vaccination against infections.

###### Long Term Care Facilities

There were four national LTCFs studies,^17–20^ two regional studies^21, 22^ and one city-wide study.^23^ Five of the seven LTCF surveillance studies utilized RT-PCR and or rapid antigen testing.^18, 20–23^

The frequency of testing appeared to vary depending on whether there was an outbreak. In one study involving LTCFs in Catalonia, Spain, the public health guidelines required routine two-to-four weekly screening of residents and staff with RT-PCR or antigen tests,^21^ another study reported two weekly screenings in the absence of an outbreak with an additional daily rapid antigen testing (with confirmatory RT-PCR for positive tests) during an outbreak.^22^ Other studies reported a more frequent screening schedule varying between five-to-seven days during an outbreak.^18, 19^ Of note, one national USA study reported that the Centres for Medicare and Medicaid services required all nursing homes in the same county to test all their staff and residents at the same frequency based on the county COVID-19 rate.^17^ A number of the LTCF studies conducted genetic sequencing to identify variants.^19, 22^ Although breakthrough cases were found among vaccinated individuals, the studies generally demonstrated the effectiveness of vaccination in the prevention of infection.

###### Community

The two community-based population surveillance studies identified in this review were national studies and were conducted in Israel and England.^24, 25^ The English study utilized self-administered throat and nose swabs and questionnaire data from a random sample of the population ages 5 years and above. From this study, it was observed that 92.3% of infections in England at the time were from the B.1.1.7 lineage and 7.7% from the B.1.617.2 lineage.^24^ The Israeli national study conducted surveillance in LTCFs and the community (using samples from random drive-through testing centres)^26^ and showed that the B.1.1.7 variant was 45% more transmissible than the wild-type strain in Israel. These studies identified COVID-19 cases among vaccinated individuals, and also determined the prevalence and transmissibility of variants.

###### Implementation Considerations and Limitations of Population Surveillance

The following implementation considerations for and limitations to population surveillance were identified in the reviewed studies:

1. For studies allowing discretionary access to testing, the frequency of testing tends to be highly variable from person to person.^12^
2. Antigen tests have lower sensitivity, therefore some positive cases may be missed during asymptomatic screening.^9^
3. The timing of infections relative to PCR testing and the sensitivity of the PCR tests can result in infections being missed during follow-up.^7^
4. While large sample sizes from multicentre studies increase generalizability, several studies observed that details of local variation in practices (in healthcare settings or long-term care facilities) are lost when findings from different centres are pooled and analyzed together. Therefore, the findings of such studies may not be generalizable.^19–21^ For example, the Centres for Medicare and Medicaid Services in the USA required all nursing homes to test residents and staff at the same frequency dictated by the rate of COVID-19 in the community (not in the facility).^17^
5. Teran et al noted that, with electronic databases, it was impossible to confirm whether patients with a breakthrough infection and a previous positive SARS-CoV-2 test result had a true reinfection or had prolonged shedding from the previous infection.^23^
6. In a study with prolonged follow up period, the authors observed a progressive decline in participants’ interest.^24^

##### Genomic Surveillance

Three genomic surveillance studies were identified in this review.^27, 28^ Two were city-wide surveillance studies conducted in the USA,^27, 29^ and one was a national scale study conducted in Uruguay.^28^ All three were community-based studies. Two of the studies identified the emergence of variants in the communities. The Uruguayan national genomic surveillance study, for example, showed that variant P.1 was introduced in Uruguay at multiple times over a period of increasing mobility.^28^ Similarly, Wang et al. demonstrated the rapid emergence of L452R mutation in the San Francisco Bay Area population, with a prevalence of 24.8% in December 2020 that increased to 62.5% in March 2021.^29^

##### Wastewater surveillance

There were six wastewater surveillance studies: two were from Spain^30, 31^ and the USA^32, 33^, respectively, and one each from Canada^34^ and the UK.^35^ The frequency of wastewater sampling varied between once a week^30, 32, 35^ to three times a week.^34^

A number of benefits of wastewater surveillance were noted across studies. In Spain,^31^ wastewater surveillance predicted major outbreaks by several weeks and showed good correlation with clinical data, providing information at local level;^30^ while another study in the USA reported a rapid turnaround time (average of 1.6 days).^32^ The challenges observed included: inconsistencies in variant representation depending on where the samples were taken within the community^33^ and differences in the capacity of wastewater to predict case numbers based on the size of the wastewater treatment plants.^35^ One study reported that laboratory build-out costs approximately $100,000 USD in capital equipment and took 3 months to complete.^32^

##### Testing Air Filters on Public Buses

In one study, Hoffman et al. placed and retrieved filters in the existing air filtration systems on public buses in Seattle to test for the presence of trapped SARS-CoV-2 RNA using phenol-chloroform extraction and RT-PCR. The study detected SARS-CoV-2 RNA in 14% of public bus filters tested.^36^

##### Metagenomics

Using the nextstrain.org database, Quinonez et al. were able to predict that the rate of B.1.1.7 lineage is going to sharply decrease (near to zero) in the coming months. The authors suggested a cautious generalizability because of the lack of validation of this approach in an experimental study.^37^

##### Emerging Technologies

New surveillance technologies for COVID-19 are emerging and some of these have been validated, but not yet studied in a real-world setting. Examples include the use of wearable monitoring device to continuously monitor skin temperature, heart rate, and respiratory rate for early detection of COVID-19 symptoms^38^ and the use of deep learning-based model to detect COVID-19 infection via CT scans and chest X-rays.^39^ A number of new assay methods (Table 3) and novel sample collection methods were also identified in this review; however, not all met inclusion criteria and were therefore not included in the broader synthesis. For example, Reeves et al. described a composite autosampler that withdraws samples from wastewater outfall within surface-accessible manholes that can be used to monitor and detect SARS-CoV-2 in individual buildings and communities.^40^

**Table 3:** Emerging Technologies

| Author<br>(Month, Year) | Type of<br>Technology | Description | Efficacy |
| --- | --- | --- | --- |
| **Emerging Technology** |  |  |  |
| Hoffman et al.<sup>36</sup><br>(June, 2021)*§ | Air filter detection of SARS-CoV-2 via buses | Novel detection using air filters on 15 buses to test for the presence of trapped SARS-CoV-2 RNA between August 2020 to March 2021 in Seattle. | Presence of SARS-CoV-2 RNA was detected in 24% (20/82) samples of public bus filters. |
| Reeves et al.<sup>40</sup><br>(May 2021) | Composite autosampler of wastewater | Withdraws samples from wastewater outfall within surface-accessible manholes. It can be used to monitor and detect SARS-CoV-2 in individual buildings/small groups of buildings. | The composite sampler generally performed well; the design achieved the objectives and provided an economical sampling unit. |
| Saha et al.<sup>39</sup><br>(April, 2021) | Deep learning-based model to detect COVID-19 infection via CT scans and chest x-rays (GraphCovidNet) | A deep learning model (also known as artificial intelligence or machine-learning) that can map CT images and chest x-rays and screen for COVID-19. | The model showed an accuracy of 99% for all datasets analyzed. |
| Wendel et al.<sup>38</sup><br>(2021) | Wearable tech monitoring device (BioButton) | A wearable device that continuously monitors skin temperature, heart rate, and respiratory rate. It can be used for early detection of COVID-19 symptoms. | Pilot study to evaluate feasibility of a wearable medical-grade monitoring device. |
| **Emerging Laboratory Testing Technology** |  |  |  |
| Narasimhan et al.<sup>42</sup> (April, 2021)§ | Serological assay to identify IgM and IgG spike protein in infected and vaccinated individuals | Used the Abbott Alinity serological assays (IgMSP and IgGSP) in combination with Abbott Alinity IgG nucleocapsid antibody test (IgGNC). The serological assay has some utility in evaluating and in distinguishing between infection and vaccination. | 100% specific for detecting SARS-CoV-2 naïve individuals. IgMSP/IgGSp were 96% and 98% sensitive for detecting inpatients with SARS-CoV-2 infection. Vaccination resulted in an increase in IgGSP/IgMSP values, with a rise in IgGSP following the second dose in the naïve group. SARS-CoV-2 infection-recovered individuals had several-fold higher IgGSP responses than naïve. |
| Stromer et al.<sup>43</sup> (Dec 2020) | Point-of-care test for rapid detection of COVID-19 antigen (NADAL COVID-19 Ag Test) | Point-of-care used to detect SARS-CoV-2 viral load and identify infectious individuals in a timely manner. | The point-of-care test reliably detected SARS-CoV-2 loads and rapidly identified infectious individuals. Additionally, the test can be useful to help identify patients who no longer transmit the virus. |
| Suhandynata<sup>44</sup> (March, 2021)§ | Serological assay to detect previous COVID infection or vaccination | Use of the serological assay Roche S-(spike) antibody and Diazyme neutralizing antibodies (NAbs) assay to detect between SARS-CoV-2 infected and vaccinated individuals. | A combination of S-antibody and N-antibody assays can be used to differentiate naturally infected individuals from vaccinated individuals, as naturally infected individuals are positive on both the S and N-antibody assays. |
| Trick et al.<sup>45</sup> (May 2021)§ | Magnetofluidic cartridge platform | Magnetofluidic cartridge platform for automated PCR testing in <30 min. The cartridges were designed for multiplex detection of SARS-CoV-2 with either distinctive variant mutations or with Influenza A and B. | The platform had a limit of detection of 2 copies/μL SARS-CoV-2 RNA with successful identification of B.1.1.7 and B.1.351 variants. Testing with nasopharyngeal swab eluates showed high sensitivity/specificity of SARS-CoV-2, Influenza A and influenza B. Further testing with saliva demonstrated successful detection of all SARS-CoV-2 positive samples with no false positives. |
| Wang et al.<sup>46</sup> (May, 2021)* | PCR screening with new assay design for VOI/VOC (multiplex allele specific RT-qPCR) | A genotyping RT-qPCR to conduct high-throughput SARS-CoV-2 variant screening as an alternative to whole-genome sequencing. | The assay had 100% concordance with whole-genome sequencing in a validation subset of 229 specimens, and was able to detect B.1.1.7, B.1.351, B.1.427, B.1.429, B.1.526, and P.2 variants (among others). |

Studies marked with an asterisk (\*) were included in the review of scientific evidence; more information on their findings can be found in Table 2.
§ Indicates preprint.
Abbreviations: RT-PCR: reverse transcriptase polymerase chain reaction; VOC: variants of concern; VOI: variants of interest

### Discussion

This review suggests that population surveillance with PCR and or rapid antigen tests were the most commonly used surveillance methods. Other approaches include: genomic surveillance, wastewater surveillance, metagenomics, and sampling of filters on public transport.

The RT-PCR detects the RNA genome of SARS-CoV-2 and has been the mainstay of COVID-19 diagnosis.^47^ As observed in several studies in this review, rapid antigen testing was often used complementarily with RT-PCR and rarely alone as a surveillance tool. This test detects the presence of viral proteins,^47^ is easy to perform, and can be interpreted without specialized training or equipment, thus can be widely distributed with a rapid turnaround time between sampling and results. Rapid antigen tests have generally relatively lower sensitivities compared with RT-PCR.^47–49^ Consequently, the European Centres for Disease Control (ECDC) suggested a more nuanced approach to rapid antigen testing, suggesting that in a high prevalence setting, a positive result from an antigen test is likely to indicate a true infection and may not require confirmation by RT-PCR;^50^ while any negative test result should be confirmed by RT-PCR immediately.^50^ Conversely, the ECDC suggests that in a low prevalence setting, rapid antigen tests should be able to rule out a highly infectious case; as such, a negative test result may not require confirmation by RT-PCR, whereas a positive test will need immediate sampling for a confirmation by RT-PCR.^50^

Population-level tracking of the origin, distribution, and trends of Covid-19 is challenging, especially considering the rapidly evolving profile of the virus. Wastewater surveillance may provide a non-invasive, anonymous and scalable method of tracking the virus within the population, within a geographic area, at a point in time.^51^ However, challenges such as inconsistencies in variant representation,^33^ differences in the capacity of wastewater to predict cases^35^ and build-out cost^32^ were identified in this review.

A limitation of this review is the lack of details on the methodological approaches to COVID-19 surveillance in the included studies. The majority of the studies were designed as epidemiological studies of existing surveillance programs, therefore, the authors focused on the prespecified study outcomes rather than the practicality of the surveillance programs. Although all the studies included vaccinated populations, there were variations in the reporting of vaccination rate. While some small hospital-based and LTCF studies reported institutional vaccination rates, several large studies did not.

## Section 2: International Guidance on Surveillance Methods in a Vaccinated Population

### Methods

Due to the anticipation that the primary evidence would stem from websites of international government organizations, a database search was not conducted. A grey literature search was conducted, including a thorough search of Google, websites of international government organizations (e.g., Center for Disease Control and Prevention [CDC], World Health Organization [WHO]), and McMaster Health Forum (COVID-END). This search was primarily examining surveillance guidance published since December 2020 (to capture guidance specific to vaccinations); however, it was expanded to include guidance on surveillance programs that would have been established prior to December 2020 but were still in place. There were no language limitations.

A screening form based on the eligibility criteria was prepared. Citations identified as potentially relevant from the literature search were screened by single reviewer across a team of four reviewers and subsequently read in full text by two reviewers and assessed for eligibility based on the criteria outlined below (Table 4). Discrepancies were resolved by discussion or by a third reviewer. Reference lists of included studies were hand searched to ensure all relevant literature is captured.

**Table 4:** Criteria for Inclusion of International Guidance on Surveillance

|  |  |
| --- | --- |
| Population | Persons who have been partially or fully vaccinated against COVID-19; populations in settings with a high vaccination rate/low prevalence rate of COVID-19 and low vaccination rate/low prevalence rate of COVID-19 were also considered |
| Intervention | Surveillance approaches to monitor for resurgence of COVID-19 and variants of concern (e.g., wastewater surveillance and metagenomics, population screening with rapid antigen testing) |
| Outcome | Any |
| Guideline Body | Guidance issued by international health organizations |
| Publication Year | Limited to publication date December 2020-onwards; ongoing surveillance programs established before December 2020 were considered for inclusion |

A standardized data extraction sheet was used to extract the month and year of publication, country, scope (e.g., national), surveillance method, vaccination status, population, setting, intended outcomes (e.g., variant surveillance), platform used for surveillance (e.g., any database), guidance summary, and implementation considerations. All reviewers completed a calibration exercise whereby data from two sample studies were extracted by all four reviewers and areas of disagreement were discussed. Data were extracted by one reviewer.

A high-level summary of the guidance pertaining to surveillance across different countries is presented below, followed by a brief discussion of the evidence included in this review; given the rapid nature of this request, a formal risk of bias assessment was not conducted. A patient partner was engaged during the co-production of a plain language summary, which is presented in a separate document.

### Results

Through hand searching of grey literature, 68 guidance documents were captured and screened for eligibility. After full-text review, a total of 42 guideline documents were excluded. The most common reason for exclusion was publication date prior to December 2020, without clear indication the surveillance methods were ongoing (n=18). Other reasons for exclusion were: not a guidance document (n=15), not surveillance method(s) (n=6), or duplicate (n=3). A total of 26 guidance documents were included in the synthesis (Table 5); see Table 6 in Appendix B for a summary of the guidance provided across included documents. Most were not specific to vaccinated populations but reported on a surveillance method of COVID-19 and were therefore included in the review; it was assumed that they were still in effect but have not yet updated.

**Table 5:**
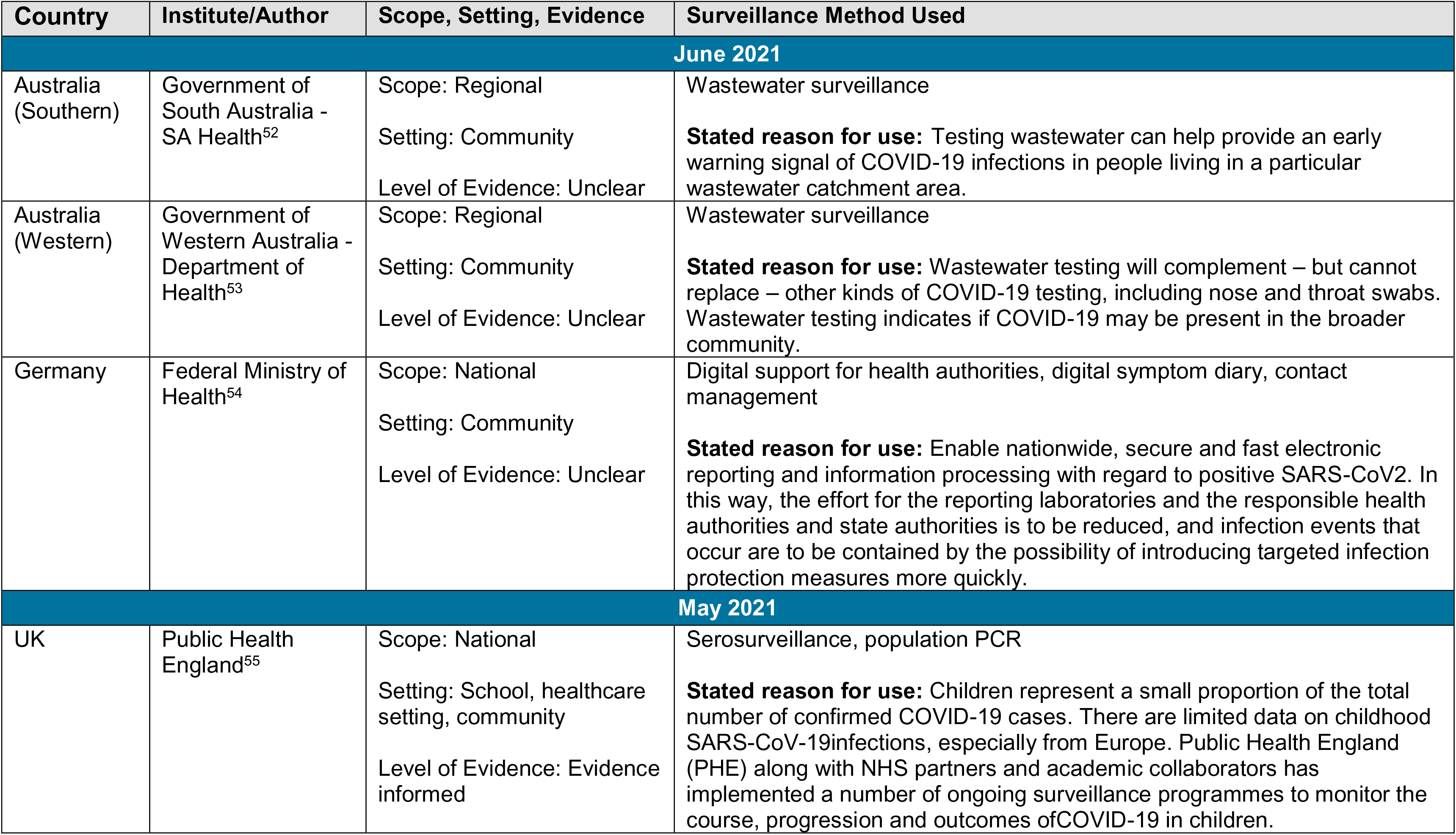
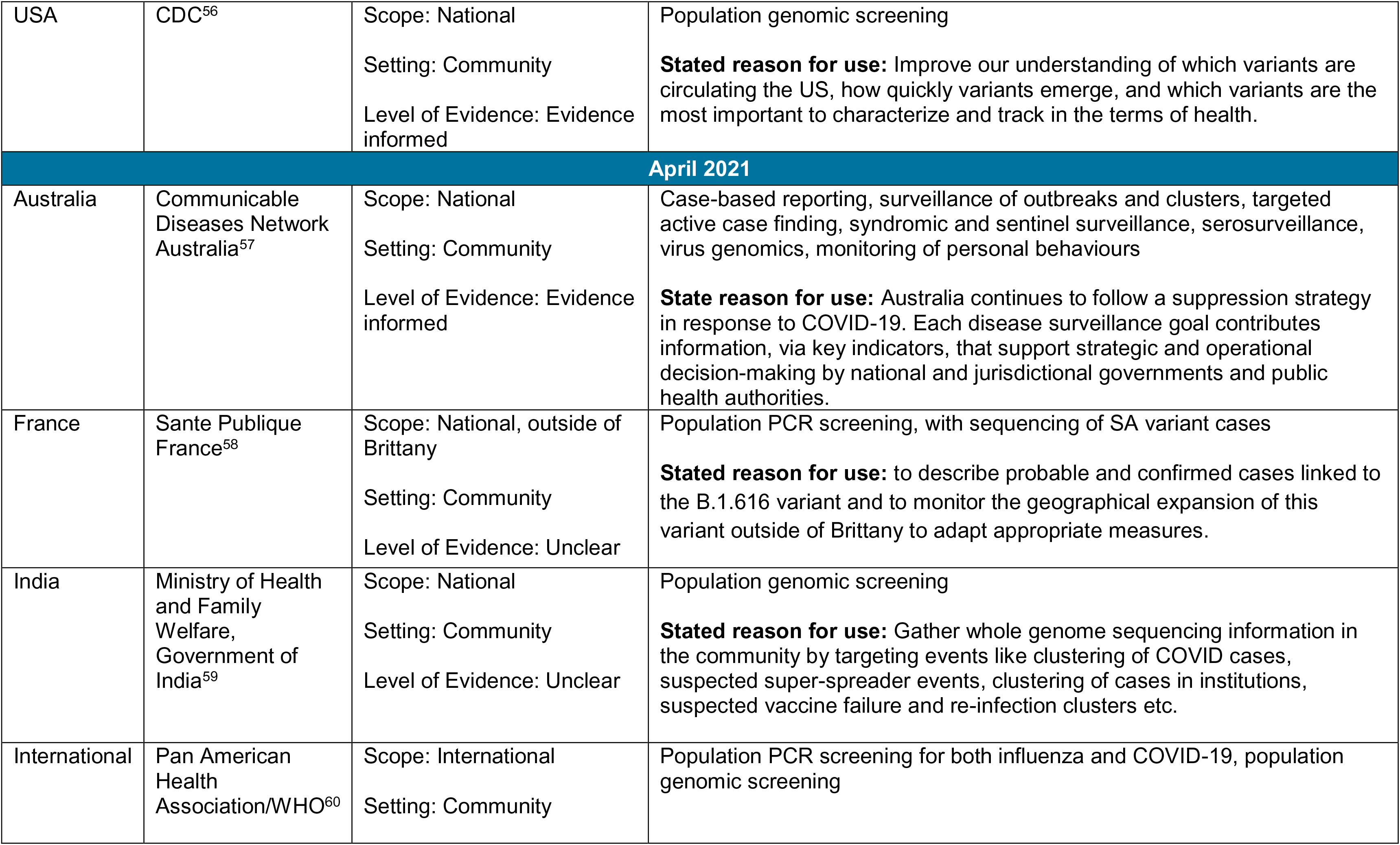
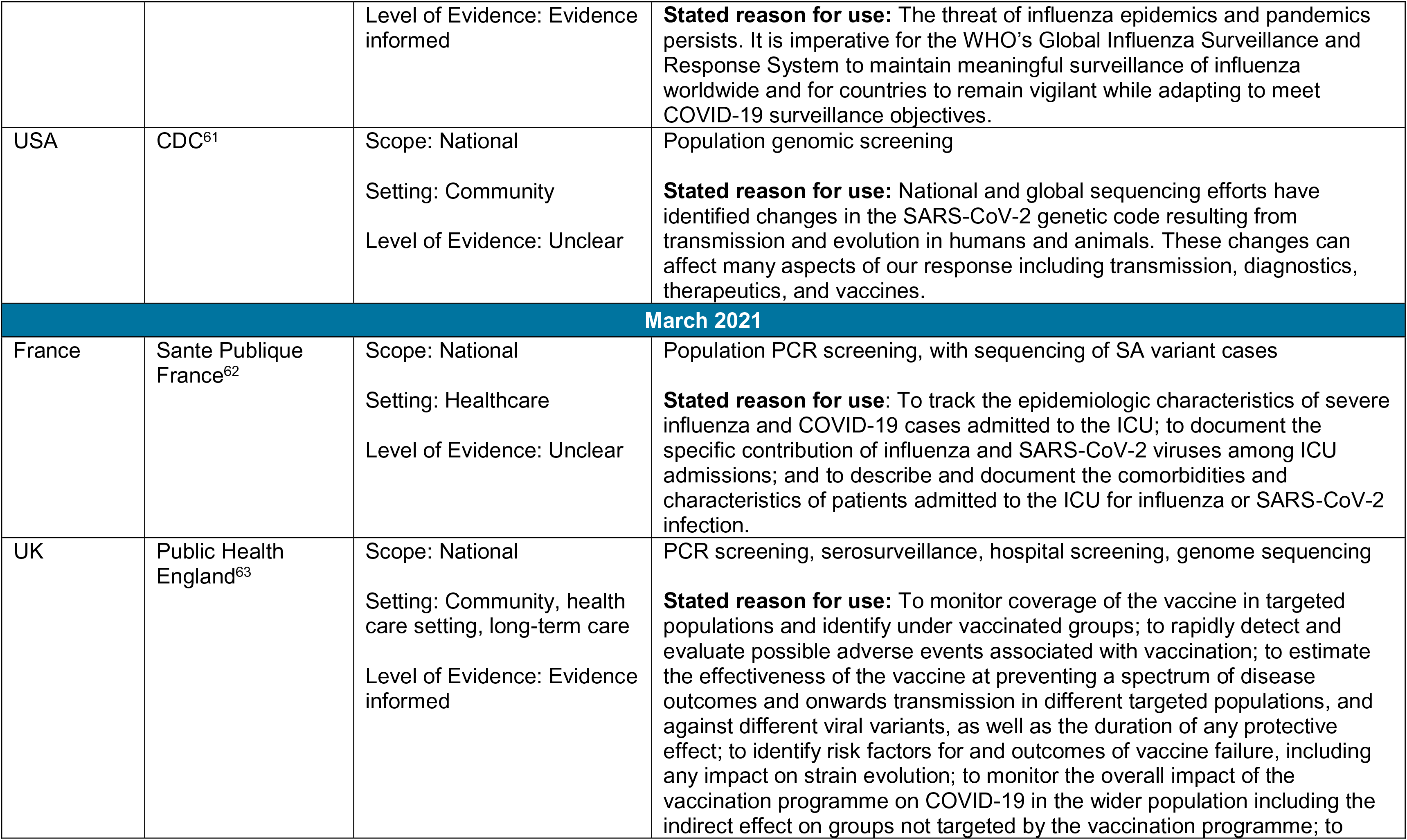
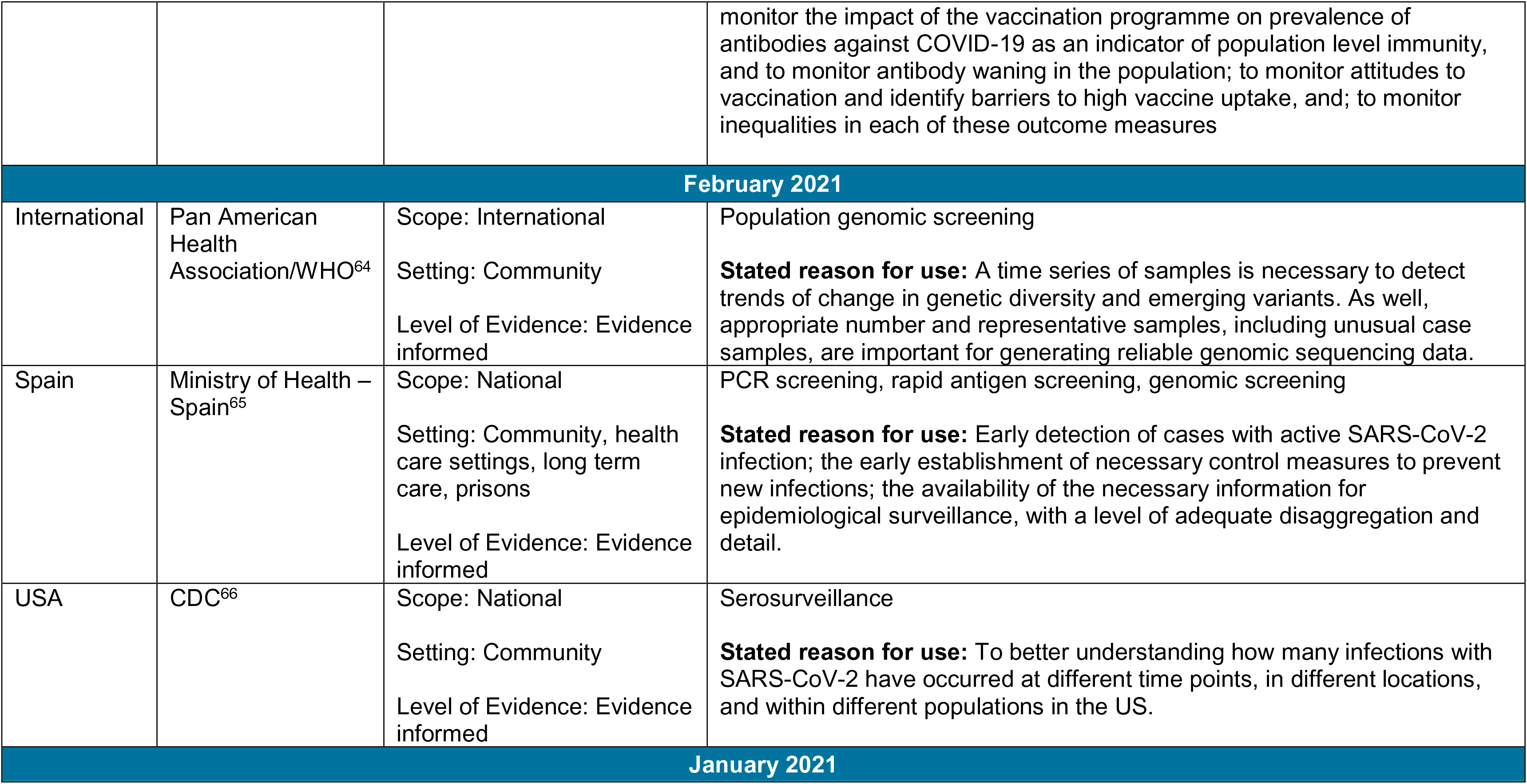
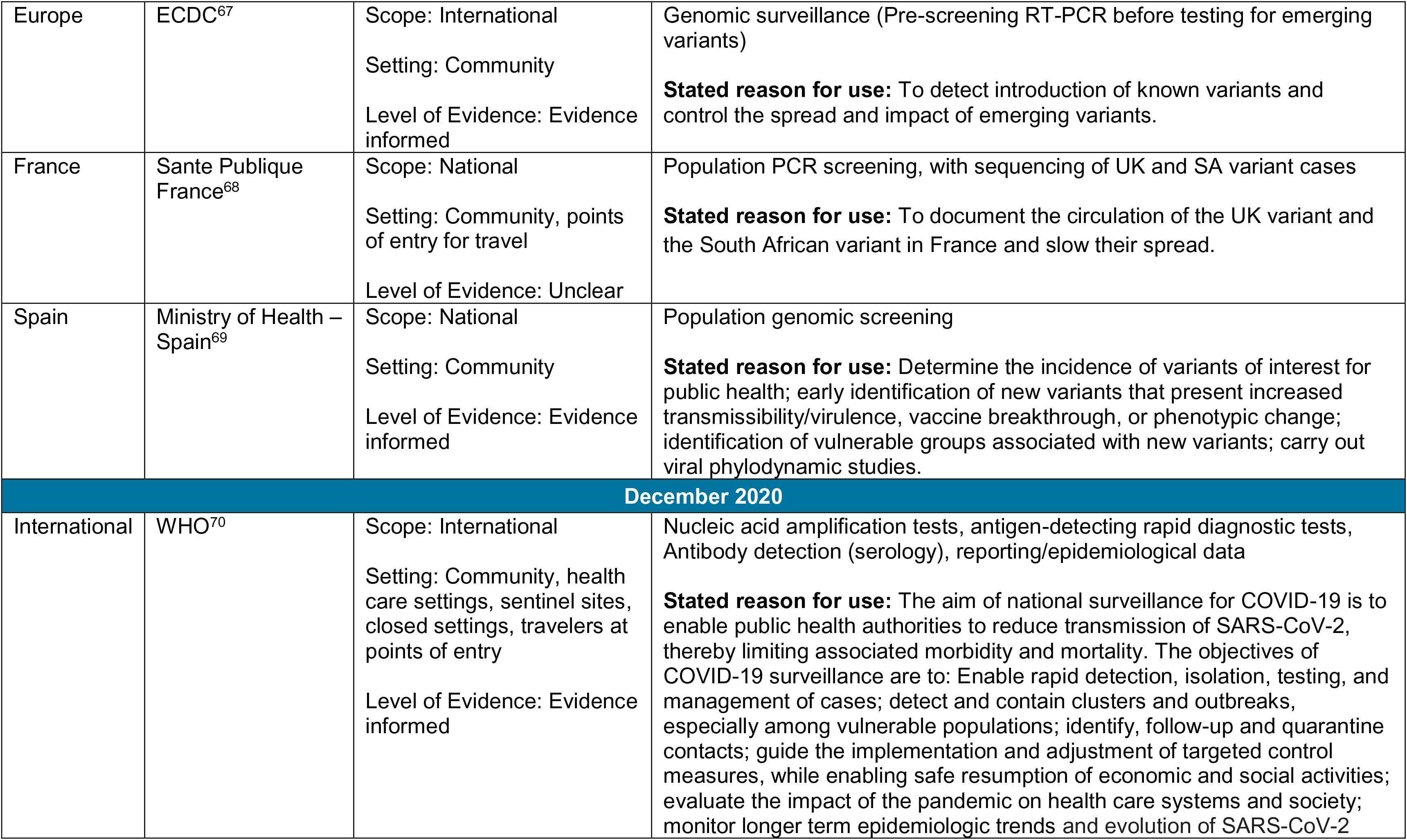
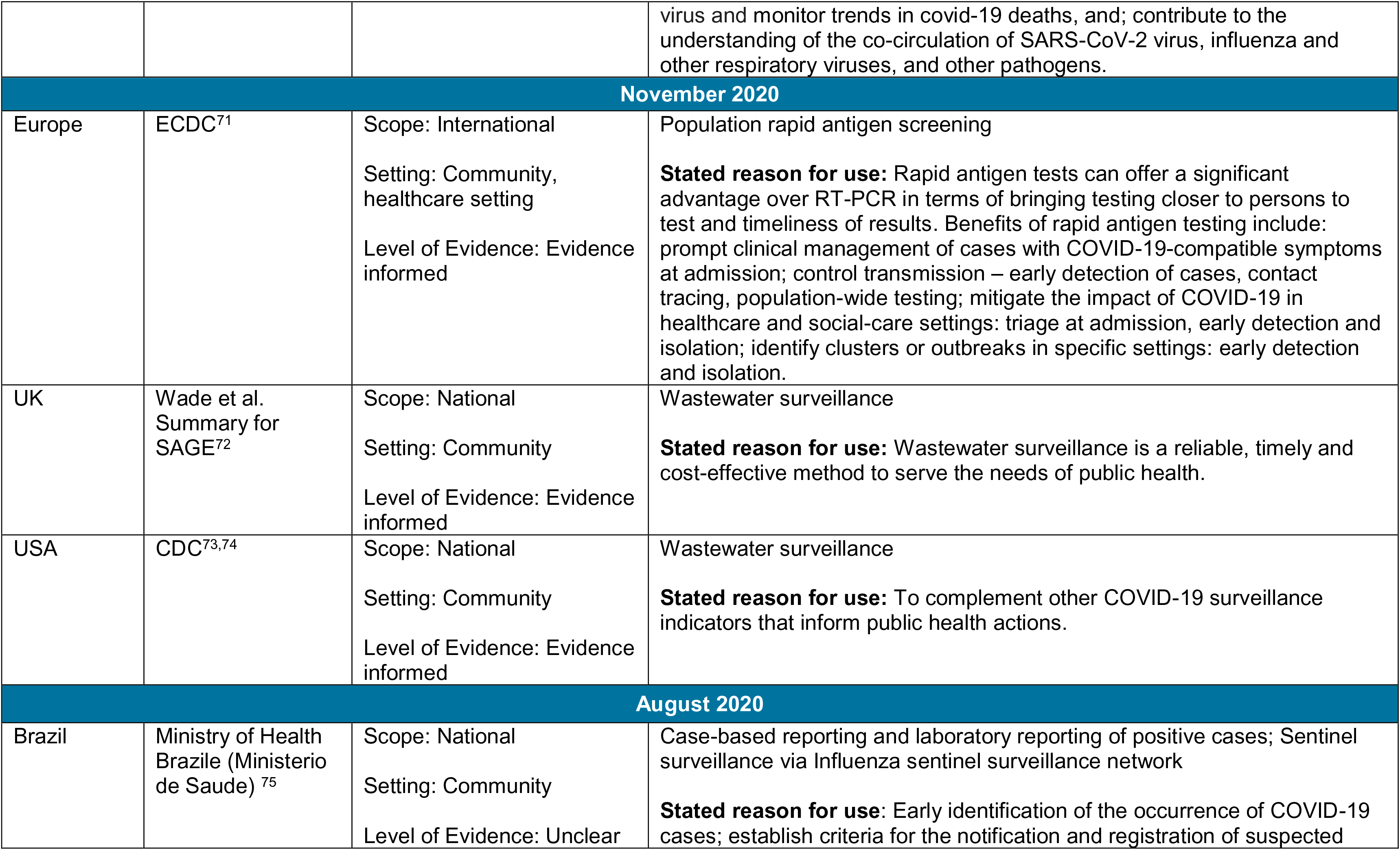
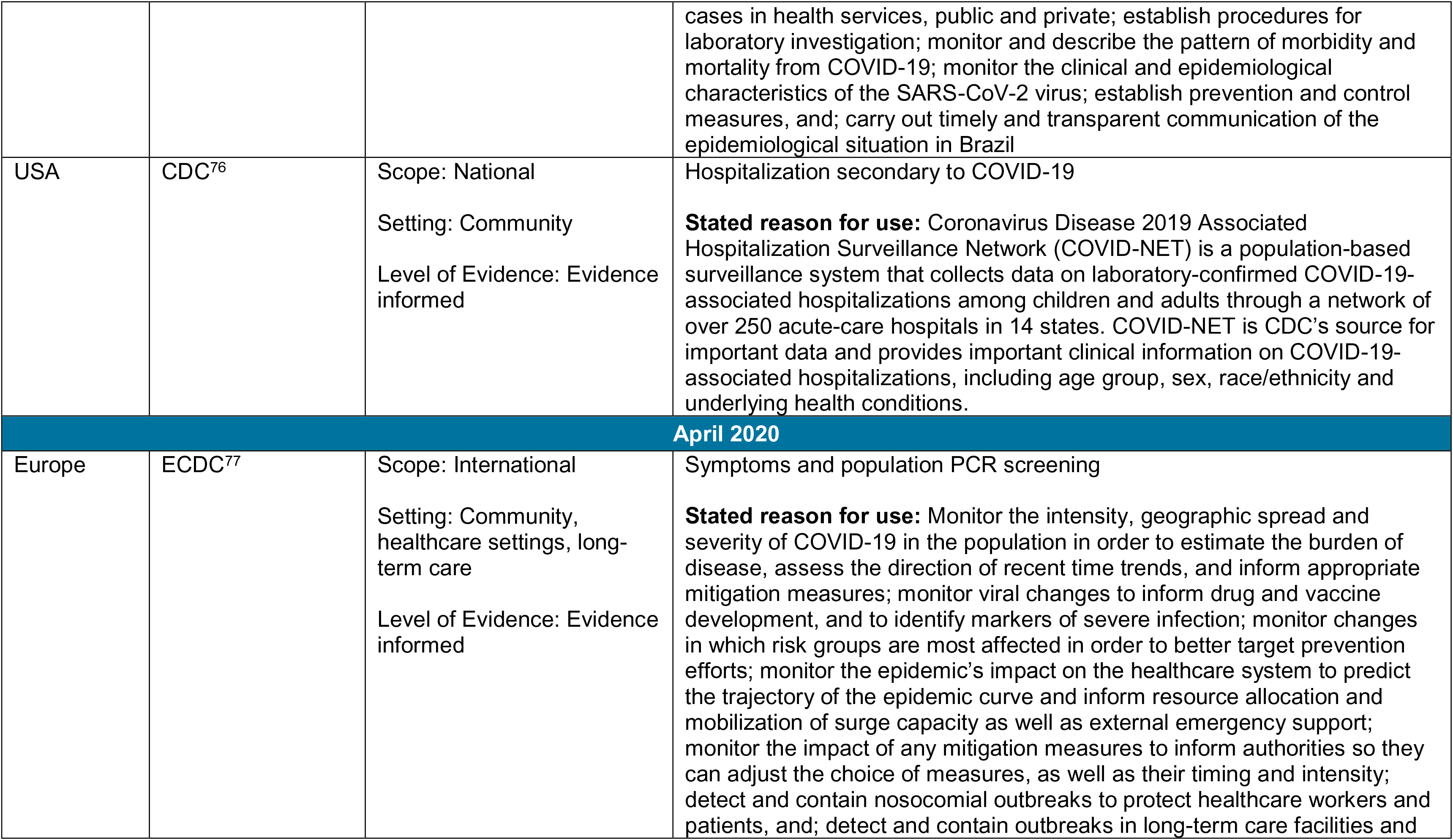
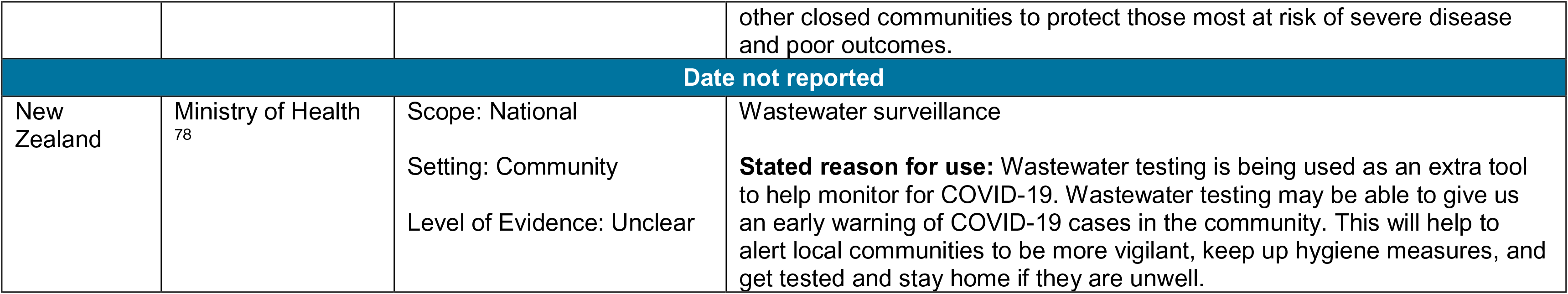
Summary of International Guidance, by Date of Publication

| Country | Institute/Author | Scope, Setting, Evidence | Surveillance Method Used |
| --- | --- | --- | --- |
| **June 2021** |  |  |  |
| Australia (Southern) | Government of South Australia - SA Health<sup>52</sup> | Scope: Regional<br>Setting: Community<br>Level of Evidence: Unclear | Wastewater surveillance<br>**Stated reason for use:** Testing wastewater can help provide an early warning signal of COVID-19 infections in people living in a particular wastewater catchment area. |
| Australia (Western) | Government of Western Australia - Department of Health<sup>53</sup> | Scope: Regional<br>Setting: Community<br>Level of Evidence: Unclear | Wastewater surveillance<br>**Stated reason for use:** Wastewater testing will complement – but cannot replace – other kinds of COVID-19 testing, including nose and throat swabs. Wastewater testing indicates if COVID-19 may be present in the broader community. |
| Germany | Federal Ministry of Health<sup>54</sup> | Scope: National<br>Setting: Community<br>Level of Evidence: Unclear | Digital support for health authorities, digital symptom diary, contact management<br>**Stated reason for use:** Enable nationwide, secure and fast electronic reporting and information processing with regard to positive SARS-CoV2. In this way, the effort for the reporting laboratories and the responsible health authorities and state authorities is to be reduced, and infection events that occur are to be contained by the possibility of introducing targeted infection protection measures more quickly. |
| **May 2021** |  |  |  |
| UK | Public Health England<sup>55</sup> | Scope: National<br>Setting: School, healthcare setting, community<br>Level of Evidence: Evidence informed | Serosurveillance, population PCR<br>**Stated reason for use:** Children represent a small proportion of the total number of confirmed COVID-19 cases. There are limited data on childhood SARS-CoV-19infections, especially from Europe. Public Health England (PHE) along with NHS partners and academic collaborators has implemented a number of ongoing surveillance programmes to monitor the course, progression and outcomes ofCOVID-19 in children. |
| USA | CDC<sup>56</sup> | Scope: National<br>Setting: Community<br>Level of Evidence: Evidence informed | Population genomic screening<br>**Stated reason for use:** Improve our understanding of which variants are circulating the US, how quickly variants emerge, and which variants are the most important to characterize and track in the terms of health. |
| April 2021 |  |  |  |
| Australia | Communicable Diseases Network Australia<sup>57</sup> | Scope: National<br>Setting: Community<br>Level of Evidence: Evidence informed | Case-based reporting, surveillance of outbreaks and clusters, targeted active case finding, syndromic and sentinel surveillance, serosurveillance, virus genomics, monitoring of personal behaviours<br>**State reason for use:** Australia continues to follow a suppression strategy in response to COVID-19. Each disease surveillance goal contributes information, via key indicators, that support strategic and operational decision-making by national and jurisdictional governments and public health authorities. |
| France | Sante Publique France<sup>58</sup> | Scope: National, outside of Brittany<br>Setting: Community<br>Level of Evidence: Unclear | Population PCR screening, with sequencing of SA variant cases<br>**Stated reason for use:** to describe probable and confirmed cases linked to the B.1.616 variant and to monitor the geographical expansion of this variant outside of Brittany to adapt appropriate measures. |
| India | Ministry of Health and Family Welfare, Government of India<sup>59</sup> | Scope: National<br>Setting: Community<br>Level of Evidence: Unclear | Population genomic screening<br>**Stated reason for use:** Gather whole genome sequencing information in the community by targeting events like clustering of COVID cases, suspected super-spreader events, clustering of cases in institutions, suspected vaccine failure and re-infection clusters etc. |
| International | Pan American Health Association/WHO<sup>60</sup> | Scope: International<br>Setting: Community | Population PCR screening for both influenza and COVID-19, population genomic screening |
|  |  | Level of Evidence: Evidence informed | **Stated reason for use:** The threat of influenza epidemics and pandemics persists. It is imperative for the WHO's Global Influenza Surveillance and Response System to maintain meaningful surveillance of influenza worldwide and for countries to remain vigilant while adapting to meet COVID-19 surveillance objectives. |
| USA | CDC<sup>61</sup> | Scope: National<br>Setting: Community<br>Level of Evidence: Unclear | Population genomic screening<br>**Stated reason for use:** National and global sequencing efforts have identified changes in the SARS-CoV-2 genetic code resulting from transmission and evolution in humans and animals. These changes can affect many aspects of our response including transmission, diagnostics, therapeutics, and vaccines. |
| **March 2021** |  |  |  |
| France | Sante Publique France<sup>62</sup> | Scope: National<br>Setting: Healthcare<br>Level of Evidence: Unclear | Population PCR screening, with sequencing of SA variant cases<br>**Stated reason for use:** To track the epidemiologic characteristics of severe influenza and COVID-19 cases admitted to the ICU; to document the specific contribution of influenza and SARS-CoV-2 viruses among ICU admissions; and to describe and document the comorbidities and characteristics of patients admitted to the ICU for influenza or SARS-CoV-2 infection. |
| UK | Public Health England<sup>63</sup> | Scope: National<br>Setting: Community, health care setting, long-term care<br>Level of Evidence: Evidence informed | PCR screening, serosurveillance, hospital screening, genome sequencing<br>**Stated reason for use:** To monitor coverage of the vaccine in targeted populations and identify under vaccinated groups; to rapidly detect and evaluate possible adverse events associated with vaccination; to estimate the effectiveness of the vaccine at preventing a spectrum of disease outcomes and onwards transmission in different targeted populations, and against different viral variants, as well as the duration of any protective effect; to identify risk factors for and outcomes of vaccine failure, including any impact on strain evolution; to monitor the overall impact of the vaccination programme on COVID-19 in the wider population including the indirect effect on groups not targeted by the vaccination programme; to monitor the impact of the vaccination programme on prevalence of antibodies against COVID-19 as an indicator of population level immunity, and to monitor antibody waning in the population; to monitor attitudes to vaccination and identify barriers to high vaccine uptake, and; to monitor inequalities in each of these outcome measures |
| **February 2021** |  |  |  |
| International | Pan American Health Association/WHO<sup>64</sup> | Scope: International<br>Setting: Community<br>Level of Evidence: Evidence informed | Population genomic screening<br>**Stated reason for use:** A time series of samples is necessary to detect trends of change in genetic diversity and emerging variants. As well, appropriate number and representative samples, including unusual case samples, are important for generating reliable genomic sequencing data. |
| Spain | Ministry of Health – Spain<sup>65</sup> | Scope: National<br>Setting: Community, health care settings, long term care, prisons<br>Level of Evidence: Evidence informed | PCR screening, rapid antigen screening, genomic screening<br>**Stated reason for use:** Early detection of cases with active SARS-CoV-2 infection; the early establishment of necessary control measures to prevent new infections; the availability of the necessary information for epidemiological surveillance, with a level of adequate disaggregation and detail. |
| USA | CDC<sup>66</sup> | Scope: National<br>Setting: Community<br>Level of Evidence: Evidence informed | Serosurveillance<br>**Stated reason for use:** To better understanding how many infections with SARS-CoV-2 have occurred at different time points, in different locations, and within different populations in the US. |
| **January 2021** |  |  |  |
| Europe | ECDC<sup>67</sup> | Scope: International<br>Setting: Community<br>Level of Evidence: Evidence informed | Genomic surveillance (Pre-screening RT-PCR before testing for emerging variants)<br>**Stated reason for use:** To detect introduction of known variants and control the spread and impact of emerging variants. |
| France | Sante Publique France<sup>68</sup> | Scope: National<br>Setting: Community, points of entry for travel<br>Level of Evidence: Unclear | Population PCR screening, with sequencing of UK and SA variant cases<br>**Stated reason for use:** To document the circulation of the UK variant and the South African variant in France and slow their spread. |
| Spain | Ministry of Health – Spain<sup>69</sup> | Scope: National<br>Setting: Community<br>Level of Evidence: Evidence informed | Population genomic screening<br>**Stated reason for use:** Determine the incidence of variants of interest for public health; early identification of new variants that present increased transmissibility/virulence, vaccine breakthrough, or phenotypic change; identification of vulnerable groups associated with new variants; carry out viral phylodynamic studies. |
| **December 2020** |  |  |  |
| International | WHO<sup>70</sup> | Scope: International<br>Setting: Community, health care settings, sentinel sites, closed settings, travelers at points of entry<br>Level of Evidence: Evidence informed | Nucleic acid amplification tests, antigen-detecting rapid diagnostic tests, Antibody detection (serology), reporting/epidemiological data<br>**Stated reason for use:** The aim of national surveillance for COVID-19 is to enable public health authorities to reduce transmission of SARS-CoV-2, thereby limiting associated morbidity and mortality. The objectives of COVID-19 surveillance are to: Enable rapid detection, isolation, testing, and management of cases; detect and contain clusters and outbreaks, especially among vulnerable populations; identify, follow-up and quarantine contacts; guide the implementation and adjustment of targeted control measures, while enabling safe resumption of economic and social activities; evaluate the impact of the pandemic on health care systems and society; monitor longer term epidemiologic trends and evolution of SARS-CoV-2 |
|  |  |  | virus and monitor trends in covid-19 deaths, and; contribute to the understanding of the co-circulation of SARS-CoV-2 virus, influenza and other respiratory viruses, and other pathogens. |
| **November 2020** |  |  |  |
| Europe | ECDC<sup>71</sup> | Scope: International<br>Setting: Community, healthcare setting<br>Level of Evidence: Evidence informed | Population rapid antigen screening<br>**Stated reason for use:** Rapid antigen tests can offer a significant advantage over RT-PCR in terms of bringing testing closer to persons to test and timeliness of results. Benefits of rapid antigen testing include: prompt clinical management of cases with COVID-19-compatible symptoms at admission; control transmission – early detection of cases, contact tracing, population-wide testing; mitigate the impact of COVID-19 in healthcare and social-care settings: triage at admission, early detection and isolation; identify clusters or outbreaks in specific settings: early detection and isolation. |
| UK | Wade et al. Summary for SAGE<sup>72</sup> | Scope: National<br>Setting: Community<br>Level of Evidence: Evidence informed | Wastewater surveillance<br>**Stated reason for use:** Wastewater surveillance is a reliable, timely and cost-effective method to serve the needs of public health. |
| USA | CDC<sup>73,74</sup> | Scope: National<br>Setting: Community<br>Level of Evidence: Evidence informed | Wastewater surveillance<br>**Stated reason for use:** To complement other COVID-19 surveillance indicators that inform public health actions. |
| **August 2020** |  |  |  |
| Brazil | Ministry of Health Brazile (Ministerio de Saude)<sup>75</sup> | Scope: National<br>Setting: Community<br>Level of Evidence: Unclear | Case-based reporting and laboratory reporting of positive cases; Sentinel surveillance via Influenza sentinel surveillance network<br>**Stated reason for use:** Early identification of the occurrence of COVID-19 cases; establish criteria for the notification and registration of suspected cases in health services, public and private; establish procedures for laboratory investigation; monitor and describe the pattern of morbidity and mortality from COVID-19; monitor the clinical and epidemiological characteristics of the SARS-CoV-2 virus; establish prevention and control measures, and; carry out timely and transparent communication of the epidemiological situation in Brazil |
| USA | CDC<sup>76</sup> | Scope: National<br>Setting: Community<br>Level of Evidence: Evidence informed | Hospitalization secondary to COVID-19<br>**Stated reason for use:** Coronavirus Disease 2019 Associated Hospitalization Surveillance Network (COVID-NET) is a population-based surveillance system that collects data on laboratory-confirmed COVID-19-associated hospitalizations among children and adults through a network of over 250 acute-care hospitals in 14 states. COVID-NET is CDC's source for important data and provides important clinical information on COVID-19-associated hospitalizations, including age group, sex, race/ethnicity and underlying health conditions. |
| **April 2020** |  |  |  |
| Europe | ECDC<sup>77</sup> | Scope: International<br>Setting: Community, healthcare settings, long-term care<br>Level of Evidence: Evidence informed | Symptoms and population PCR screening<br>**Stated reason for use:** Monitor the intensity, geographic spread and severity of COVID-19 in the population in order to estimate the burden of disease, assess the direction of recent time trends, and inform appropriate mitigation measures; monitor viral changes to inform drug and vaccine development, and to identify markers of severe infection; monitor changes in which risk groups are most affected in order to better target prevention efforts; monitor the epidemic's impact on the healthcare system to predict the trajectory of the epidemic curve and inform resource allocation and mobilization of surge capacity as well as external emergency support; monitor the impact of any mitigation measures to inform authorities so they can adjust the choice of measures, as well as their timing and intensity; detect and contain nosocomial outbreaks to protect healthcare workers and patients, and; detect and contain outbreaks in long-term care facilities and other closed communities to protect those most at risk of severe disease and poor outcomes. |
| **Date not reported** |  |  |  |
| New Zealand | Ministry of Health<sup>78</sup> | Scope: National<br>Setting: Community<br>Level of Evidence: Unclear | Wastewater surveillance<br>**Stated reason for use:** Wastewater testing is being used as an extra tool to help monitor for COVID-19. Wastewater testing may be able to give us an early warning of COVID-19 cases in the community. This will help to alert local communities to be more vigilant, keep up hygiene measures, and get tested and stay home if they are unwell. |

Guidance documents from 11 countries/regions were identified including Australia,^52, 53, 57^ Brazil,^75^ Europe,^67, 71, 77^ France,^58, 62, 68^ Germany,^54^ India,^59^ International,^60, 64, 70^ New Zealand,^78^ Spain,^65, 69^ the United Kingdom^55, 63, 72^ and the United States.^56, 61, 66, 73, 74, 76^ Documents were derived from government websites (Departments/Ministries of Health, National Governments), subsidiaries of national governments (e.g., Public Health England, Centre for Disease Control), or from international organizations (e.g., World Health Organization, Pan American Health Association, and European Centre for Disease Control). The scope of the guidance documents was mostly national-focused (n=17), however there were some that were international- (n=6) and regional-focused (n=3).

All of the guidance documents included surveillance methods conducive for community settings. Other settings of interest were healthcare setting including hospitals and primary care centres,^55, 63, 65, 68, 70^ long-term care facilities,^63, 65^ points of entry for travel,^68, 70^ schools,^55^ and other sentinel sites (e.g., prisons and closed settings).^65, 70^

Seven overarching surveillance methods emerged in the literature. PCR-testing was the most recommended surveillance method (n=11), followed by genomic screening (n=9), serosurveillance (n=5), wastewater surveillance (n=5), antigen testing (n=3), health record screening (n=2), and syndromic surveillance (n=2) (Figure 3).

**Figure 3:**
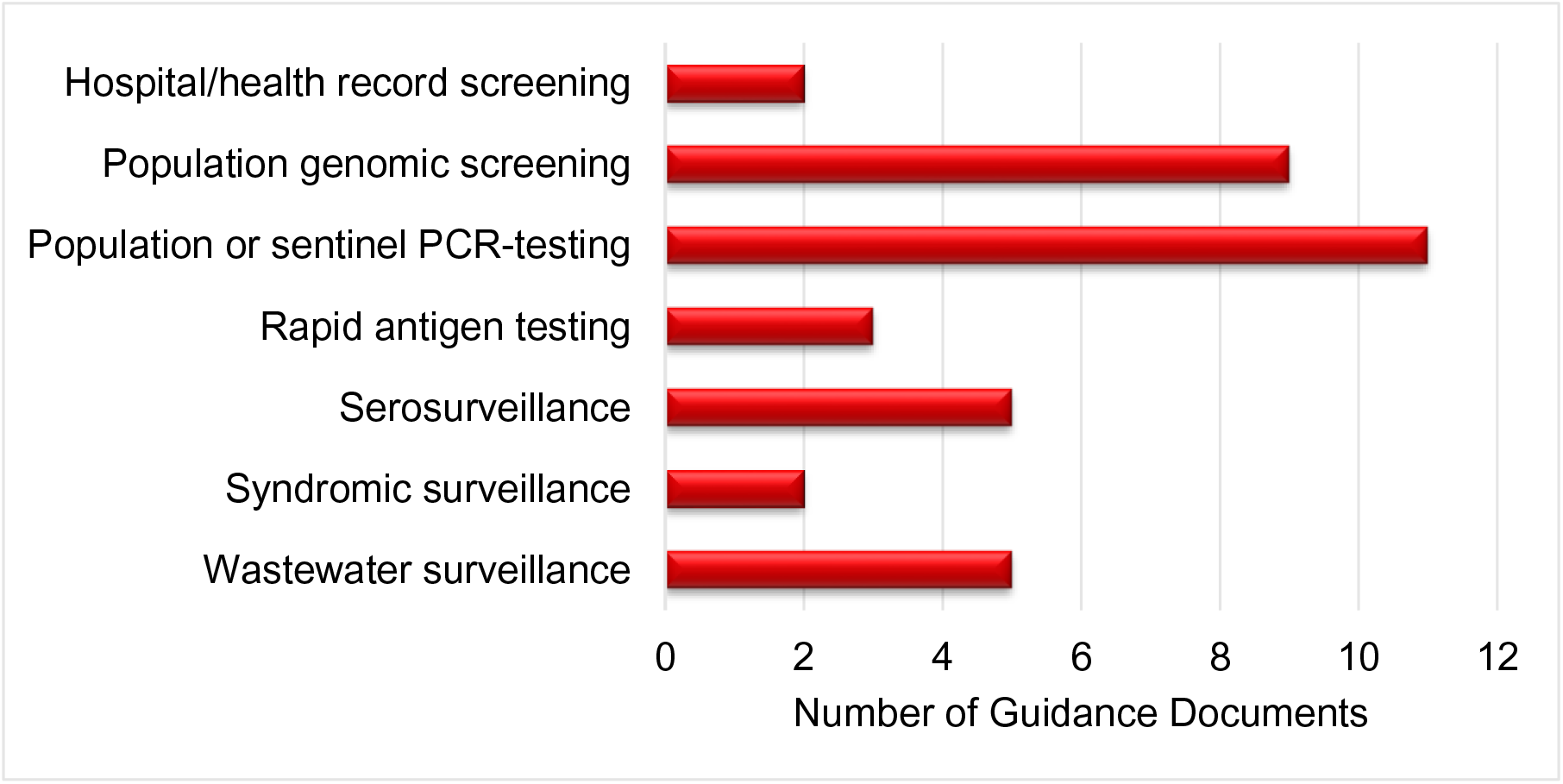
Surveillance Methods Reported in Included International Guidelines (n=26)

#### Vaccine-Specific Surveillance

Only one document was specific to surveillance methods to be deployed in a vaccinated population. The Public Health England *COVID-19 Vaccine Surveillance Strategy*^63^ recommends surveillance methods including PCR-testing, health record screening, serosurveillance, and genome sequencing with the following objectives:

- To monitor coverage of the vaccine in targeted populations and identify under-vaccinated groups;
- to rapidly detect and evaluate possible adverse events associated with vaccination;
- to estimate the effectiveness of the vaccine at preventing a spectrum of disease outcomes and onwards transmission in different targeted populations, and against different viral variants, as well as the duration of any protective effect;
- to identify risk factors for and outcomes of vaccine failure, including any impact on strain evolution;
- to monitor the overall impact of the vaccination program on COVID-19 in the wider population including the indirect effect on groups not targeted by the vaccination program;
- to monitor the impact of the vaccination program on prevalence of antibodies against COVID-19 as an indicator of population level immunity, and to monitor antibody waning in the population;
- to monitor attitudes to vaccination and identify barriers to high vaccine uptake, and;
- to monitor inequalities in each of these outcome measures.

### Discussion

Comprehensive hand-searching for international guidance on surveillance methods of COVID-19 yielded 26 documents. Most were not specific to vaccinated populations but reported on a surveillance method of COVID-19 and were therefore included in the review; it was assumed that they were still in effect but have not yet been updated. Seven surveillance methods emerged from the guidance documents: PCR-testing, genomic screening, serosurveillance, wastewater surveillance, antigen testing, health record screening, and syndromic surveillance. Many of the surveillance methods were recommended for use in community settings, however PCR-testing, antigen testing, genomic screening, serosurveillance, and health record screening were also recommended for targeted settings such as health care facilities, long-term care facilities, schools, and points of entry for travelers.

The objectives of the surveillance methods were consistent across countries. PCR-testing, antigen-testing, syndromic surveillance, health record screening, and serosurveillance should be used to: monitor the intensity, spread, and severity of COVID-19 in order to estimate the burden of disease, identify at-risk populations, identify outbreaks, to adjust public health measures as needed. Additionally, genomic sequencing should be used to identify variations and evolution of SARS-CoV-2 to identify variants of concern. Wastewater surveillance should be used to complement other surveillance methods, to detect if COVID-19 and its variants are present in a community setting.

Only one document (published by Public Health England) was identified that provided guidance specific to surveillance of vaccinated populations. Public Health England details their plan to surveil and monitor COVID-19 in vaccinated populations in the UK, including conducting cohort studies such as SIREN, VIVALDI, and CONSENSUS studies.^63^ These will involve routine surveillance, enhanced surveillance, use of electronic health records, surveillance of vaccine failure (including follow-up with viral whole genome sequencing) and sero-surveillance (including blood donor samples, routine blood tests, and residual sera).

There are several limitations to this review. Given the rapid nature of this report and the evidence of interest (i.e., international guidance), we were unable to carry-out an exhaustive systematic search of the literature. Therefore, the international guidance captured here may not include all countries’/institutes’ guidance for surveillance. Additionally, we were unable to address the quality of evidence reported in the guidance documents because of the variation in reporting and level of detail in the included documents.

## Conclusions

Evidence for post-vaccination COVID-19 surveillance was derived from studies in partially or fully vaccinated populations. Population PCR screening, supplemented by rapid antigen tests, was the most frequently used surveillance method and also the most commonly recommended across jurisdictions. The selection of testing method and the frequency of testing was determined by the intensity of the disease and the scale of testing. Other common testing methods included wastewater surveillance and genomic surveillance. A few novel technologies are emerging, however, many of these are yet to be utilized in the real-world setting. There is limited evidence-based guidance on surveillance in a vaccinated population. Most recent guidance on COVID-19 surveillance is not specific to vaccinated individuals, or it is in effect but has not yet been updated to reflect that. Therefore, more evidence-informed guidance on testing and surveillance approaches in a vaccinated population that incorporates all testing modalities is required.

## Data Availability

All datasets supporting the conclusions of this article are included within the article

## Funding Acknowledgement(s)

The SPOR Evidence Alliance (<u>SPOR EA</u>) is supported by the Canadian Institutes of Health Research (<u>CIHR</u>) under the Strategy for Patient-Oriented Research (<u>SPOR</u>) initiative.

COVID-19 Evidence Network to support Decision-making (<u>COVID-END</u>) is supported by the Canadian Institutes of Health Research (<u>CIHR</u>) through the Canadian 2019 Novel Coronavirus (COVID-19) Rapid Research Funding opportunity.

## Third-Party Materials

If you wish to reuse non-textual material from this report that is attributed to a third party, such as tables, figures or images, it is your responsibility to determine whether permission is required for such use and to obtain necessary permission from the copyright holder. The risk of claims resulting from infringement of any third-party-owned material rests solely with the user.

## General Disclaimer

This report was prepared by the University of Calgary Health Technology Assessment Unit on behalf of the SPOR Evidence Alliance and COVID-END. It was developed through the analysis, interpretation and synthesis of scientific research and/or health technology assessments published in peer-reviewed journals, institutional websites and other distribution channels. It also incorporates selected information provided by experts and patient/citizen partners with lived experience on the subject matter. This document may not fully reflect all the scientific evidence available at the time this report was prepared. Other relevant scientific findings may have been reported since completion of this synthesis report.

SPOR Evidence Alliance, COVID-END and the project team make no warranty, express or implied, nor assume any legal liability or responsibility for the accuracy, completeness, or usefulness of any information, data, product, or process disclosed in this report. Conclusions drawn from, or actions undertaken on the basis of, information included in this report are the sole responsibility of the user.

## Abbreviations

CDC: Centres for Disease Control and Prevention
COVID-19: Coronavirus Disease 2019
VOC: Variant of concern
WHO: World Health Organization
RT-PCR: Reverse transcriptase polymerase chain reaction
ECDC: European Centre for Disease Prevention and Control

## Appendix A: Search Strategies for Scientific Evidence of Surveillance

### Ovid Multifile

Database: EBM Reviews - Cochrane Central Register of Controlled Trials <MAY 2021>, EBM Reviews - Cochrane Database of Systematic Reviews <2005 to June 9, 2021>, Embase <1974 to 2021 June 11>, Ovid MEDLINE(R) and Epub Ahead of Print, In-Process, In-Data-Review & Other Non-Indexed Citations and Daily <1946 to June 11, 2021>

### Search Strategy

1. COVID-19/ (85559)
2. SARS-CoV-2/ (81242)
3. Coronavirus/ (13278)
4. Betacoronavirus/ (40986)
5. Coronavirus Infections/ (57260)
6. COVID-19 or COVID19).tw,kf. (253916)
7. ((coronavirus* or corona virus*) and (hubei or wuhan or beijing or shanghai)).tw,kf. (9779)
8. (wuhan adj5 virus*).tw,kf. (519)
9. 2019-nCoV or 19nCoV or 2019nCoV).tw,kf. (3166)
10. (nCoV or n-CoV or “CoV 2” or CoV2).tw,kf. (92137)
11. (SARS-CoV-2 or SARS-CoV2 or SARSCoV-2 or SARSCoV2 or SARS2 or SARS-2 or severe acute respiratory syndrome coronavirus 2).tw,kf. (93701)
12. (2019-novel CoV or Sars-coronavirus2 or Sars-coronavirus-2 or SARS-like coronavirus* or ((novel or new or nouveau) adj2 (CoV or nCoV or covid or coronavirus* or corona virus or Pandemi*2)) or (coronavirus* and pneumonia)).tw,kf. (35999)
13. (novel coronavirus* or novel corona virus* or novel CoV).tw,kf. (18244)
14. ((coronavirus* or corona virus*) adj2 “2019”).tw,kf. (58052)
15. ((coronavirus* or corona virus*) adj2 “19”).tw,kf. (9366)
16. (”coronavirus 2” or “corona virus 2”).tw,kf. (29952)
17. (OC43 or NL63 or 229E or HKU1 or HCoV* or Sars-coronavirus*).tw,kf. (7566)
18. COVID-19.rx,px,ox. or severe acute respiratory syndrome coronavirus 2.os. (6367)
19. (coronavirus* or corona virus*).ti. (43970)
20. (”B.1.1.7” or “B.1.351” or “B.1.617” or “B.1.427” or “B.1.429”).tw,kf,rx,px,ox. (626)
21. (”P.1” and (Brazil* or variant?)).tw,kf,rx,px,ox. (3431)
22. ((alpha or beta or delta or gamma) adj3 variant?).tw,kf. (11593)
23. or/1-22 [COVID-19] (329554)
24. vaccinated.tw,kf. (97271)
25. inoculated.tw,kf. (153237)
26. immuni#ed.tw,kf. (119849)
27. post-vaccinat*.tw,kf. (11845)
28. post-inoculat*.tw,kf. (11629)
29. post-immuni*.tw,kf. (3943)
30. ((after or already or full or fully or post or received) adj3 (immunis* or immuniz* or immunity or inoculat* or vaccin*)).tw,kf. (185626)
31. (status* adj3 (immunis* or immuniz* or immunity or inoculat* or vaccin*)).tw,kf. (17479)
32. or/24-31 [VACCINATED] (472108)
33. and 32 (5627)
34. Health Surveys/ (249807)
35. ((health or population?) adj3 survey?).tw,kf. (175676)
36. ((disease* or pandemic*) adj3 (monitor* or survey?)).tw,kf. (54464)
37. ((COVID or COVID-19 or COVID19) adj3 (monitor* or survey?)).tw,kf. (1920)
38. ((coronavirus* or corona virus*) adj3 (monitor* or survey?)).tw,kf. (216)
39. ((2019-nCoV or nCoV or n-CoV or SARS-CoV-2 or SARS-CoV2 or SARSCoV-2 or SARSCoV2 or SARS2) adj3 (monitor* or survey?)).tw,kf. (449)
40. ((BNT162 or BNT162-01 or BNT162a1 or BNT162b1 or BNT162b2 or BNT162c2) adj3 (monitor* or survey?)).tw,kf. (3)
41. ((alpha or beta or delta or gamma) adj3 variant? adj3 (monitor* or survey*)).tw,kf. (6)
42. *Epidemiological Methods/ (6485)
43. Epidemiological Monitoring/ (10263)
44. (epidemiolog* adj3 monitor*).tw,kf. (3222)
45. Seroepidemiologic Studies/ (26333)
46. ((seroepidemiol* or sero-epidemiol*) adj3 (monitor* or survey* or study or studies)).tw,kf. (7085)
47. (seromonitor* or sero-monitor* or serological monitor*).tw,kf. (568)
48. (seroprevalen* or sero-prevalen* or serological prevalen*).tw,kf. (46051)
49. (serosurveillan* or sero-surveillan*).tw,kf. (1271)
50. (serosurvey? or sero-survey? or serological survey?).tw,kf. (8888)
51. Wastewater-Based Epidemiological Monitoring/ (2419)
52. Waste Water/ (50317)
53. (sewage* or wastewater or waste water).tw,kf. (179449)
54. Norman score?.tw,kf. (84)
55. Data Collection/ (305772)
56. ((collect* or monitor*) adj3 data).tw,kf. (1017644)
57. Public Health Practice/ (70070)
58. ((community health or public health) adj3 (practice? or activit* or endeavo?r?)).tw,kf. (15598)
59. exp Population Surveillance/ (306931)
60. surveillance*.tw,kf. (471636)
61. (biosurveillance* or bio-surveillance*).tw,kf. (688)
62. Mass Screening/ (167946)
63. screening.tw,kf. (1426501)
64. ((mass or population*) adj3 screen*).tw,kf. (56691)
65. (screen* or detect* or identif* or recogni*).ti,kf. (2180317)
66. ((early or earlier or earliest or ongoing or regular*) adj5 (screen* or detect* or identif* or recogni*)).tw,kf. (528091)
67. (case finding? or casefinding?).tw,kf. (13173)
68. Metagenomics/ (24172)
69. (ecogenomic* or eco-genomic* or metagenomic* or meta-genomic*).tw,kf. (31596)
70. ((communit* or ecologic* or environment* or population) adj3 genomic*).tw,kf. (11485)
71. COVID-19 Testing/ (5913)
72. ((COVID or COVID-19 or COVID19) adj3 test*).tw,kf. (7582)
73. ((coronavirus* or corona virus*) adj3 test*).tw,kf. (1002)
74. ((2019-nCoV or nCoV or n-CoV or SARS-CoV-2 or SARS-CoV2 or SARSCoV-2 or SARSCoV2 or SARS2) adj3 test*).tw,kf. (7627)
75. ((BNT162 or BNT162-01 or BNT162a1 or BNT162b1 or BNT162b2 or BNT162c2) adj3 test*).tw,kf. (8)
76. ((alpha or beta or delta or gamma) adj3 variant? adj3 test*).tw,kf. (41)
77. (serologic* adj3 test*).tw,kf. (50026)
78. Point-of-Care Testing/ (18292)
79. ((point-of-care or bedside? or bed side? or POC or rapid*) adj3 (assay? or immunoassay? or immuno-assay? or detect* or diagnos* or screen* or test*)).tw,kf. (225556)
80. (field adj3 (assay? or immunoassay? or immuno-assay? or detect* or diagnos* or screen* or test*)).tw,kf. (74754)
81. POCT.tw,kf. (5011)
82. (rapid adj3 (antigen* adj3 (assay? or immunoassay? or immuno-assay? or test*))).tw,kf. (3251)
83. (random* adj3 (assay? or immunoassay? or immuno-assay? or detect* or diagnos* or screen* or test*)).tw,kf. (80902)
84. (random* adj3 sampl*).tw,kf. (173755)
85. (pool* adj3 sampl*).tw,kf. (18948)
86. COVID-19/ep [epidemiology] (14412)
87. Coronavirus Infections/ep [epidemiology] (23494)
88. Incidence/ (742644)
89. Prevalence/ (1098179)
90. (incidence or prevalen*).tw,kf. (3890564)
91. or/34-90 [SURVEILLANCE - BROAD] (9638102)
92. 33 and 91 (1620)
93. exp Animals/ not Humans/ (16106641)
94. 92 not 93 [ANIMAL-ONLY REMOVED] (1213)
95. limit 94 to yr=”2020-current” (1045)
96. 95 use ppez [MEDLINE RECORDS] (557)
97. coronavirus disease 2019/ (204518)
98. severe acute respiratory syndrome coronavirus 2/ (98155)
99. Coronavirinae/ (4902)
100. Betacoronavirus/ (40986)
101. coronavirus infection/ (58134)
102. (COVID-19 or COVID19).tw,kw. (258126)
103. ((coronavirus* or corona virus*) and (hubei or wuhan or beijing or shanghai)).tw,kw. (9934)
104. (wuhan adj5 virus*).tw,kw. (542)
105. (2019-nCoV or 19nCoV or 2019nCoV).tw,kw. (3494)
106. (nCoV or n-CoV or “CoV 2” or CoV2).tw,kw. (91701)
107. (SARS-CoV-2 or SARS-CoV2 or SARSCoV-2 or SARSCoV2 or SARS2 or SARS-2 or severe acute respiratory syndrome coronavirus 2).tw,kw. (99684)
108. (2019-novel CoV or Sars-coronavirus2 or Sars-coronavirus-2 or SARS-like coronavirus* or ((novel or new or nouveau) adj2 (CoV or nCoV or covid or coronavirus* or corona virus or Pandemi*2)) or (coronavirus* and pneumonia)).tw,kw. (36564)
109. (novel coronavirus* or novel corona virus* or novel CoV).tw,kw. (18583)
110. ((coronavirus* or corona virus*) adj2 “2019”).tw,kw. (58746)
111. ((coronavirus* or corona virus*) adj2 “19”).tw,kw. (9103)
112. (”coronavirus 2” or “corona virus 2”).tw,kw. (29862)
113. (OC43 or NL63 or 229E or HKU1 or HCoV* or Sars-coronavirus*).tw,kw. (7753)
114. (coronavirus* or corona virus*).ti. (43970)
115. (”B.1.1.7” or “B.1.351” or “B.1.617” or “B.1.427” or “B.1.429”).tw,kw. (629)
116. (”P.1” and (Brazil* or variant?)).tw,kw. (3418)
117. ((alpha or beta or delta or gamma) adj3 variant?).tw,kw. (11631)
118. or/97-117 [COVID-19] (338798)
119. vaccinated.tw,kw. (97263)
120. inoculated.tw,kw. (153233)
121. immuni#ed.tw,kw. (119853)
122. post-vaccinat*.tw,kw. (11879)
123. post-inoculat*.tw,kw. (11625)
124. post-immuni*.tw,kw. (3945)
125. ((after or already or full or fully or post or received) adj3 (immunis* or immuniz* or immunity or inoculat* or vaccin*)).tw,kw. (185537)
126. (status* adj3 (immunis* or immuniz* or immunity or inoculat* or vaccin*)).tw,kw. (17522)
127. or/119-126 [VACCINATED] (472056)
128. 118 and 127 (5590)
129. health survey/ (266784)
130. ((health or population?) adj3 survey?).tw,kw. (180468)
131. disease surveillance/ (30911)
132. ((disease* or pandemic*) adj3 (monitor* or survey?)).tw,kw. (56302)
133. ((COVID or COVID-19 or COVID19) adj3 (monitor* or survey?)).tw,kw. (2268)
134. ((coronavirus* or corona virus*) adj3 (monitor* or survey?)).tw,kw. (283)
135. ((2019-nCoV or nCoV or n-CoV or SARS-CoV-2 or SARS-CoV2 or SARSCoV-2 or SARSCoV2 or SARS2) adj3 (monitor* or survey?)).tw,kw. (523)
136. ((BNT162 or BNT162-01 or BNT162a1 or BNT162b1 or BNT162b2 or BNT162c2) adj3 (monitor* or survey?)).tw,kw. (4)
137. ((alpha or beta or delta or gamma) adj3 variant? adj3 (monitor* or survey*)).tw,kw. (6)
138. epidemiological data/ (34565)
139. epidemiological monitoring/ (10263)
140. infection rate/ (33957)
141. (epidemiolog* adj3 monitor*).tw,kw. (3329)
142. seroepidemiology/ (4490)
143. ((seroepidemiol* or sero-epidemiol*) adj3 (monitor* or survey* or study or studies)).tw,kw. (7224)
144. (seromonitor* or sero-monitor* or serological monitor*).tw,kw. (575)
145. (seroprevalen* or sero-prevalen* or serological prevalen*).tw,kw. (46903)
146. (serosurveillan* or sero-surveillan*).tw,kw. (1338)
147. (serosurvey? or sero-survey? or serological survey?).tw,kw. (9023)
148. waste water/ (50317)
149. (sewage* or wastewater or waste water).tw,kw. (182355)
150. Norman score?.tw,kw. (84)
151. information processing/ (254616)
152. ((collect* or monitor*) adj3 data).tw,kw. (1017125)
153. ((community health or public health) adj3 (practice? or activit* or endeavo?r?)).tw,kw. (15937)
154. surveillance*.tw,kw. (477847)
155. (biosurveillance* or bio-surveillance*).tw,kw. (735)
156. screening/ (289977)
157. mass screening/ (167946)
158. screening.tw,kw. (1440549)
159. ((mass or population*) adj3 screen*).tw,kw. (58731)
160. (screen* or detect* or identif* or recogni*).ti. (2128877)
161. ((early or earlier or earliest or ongoing or regular*) adj5 (screen* or detect* or identif* or recogni*)).tw,kw. (529117)
162. (case finding? or casefinding?).tw,kw. (13335)
163. metagenomics/ (24172)
164. (ecogenomic* or eco-genomic* or metagenomic* or meta-genomic*).tw,kw. (32991)
165. ((communit* or ecologic* or environment* or population) adj3 genomic*).tw,kw. (11715)
166. immunoassay/ (102667)
167. antibody detection/ (44387)
168. ((COVID or COVID-19 or COVID19) adj3 test*).tw,kw. (7790)
169. ((coronavirus* or corona virus*) adj3 test*).tw,kw. (1062)
170. ((2019-nCoV or nCoV or n-CoV or SARS-CoV-2 or SARS-CoV2 or SARSCoV-2 or SARSCoV2 or SARS2) adj3 test*).tw,kw. (7742)
171. ((BNT162 or BNT162-01 or BNT162a1 or BNT162b1 or BNT162b2 or BNT162c2) adj3 test*).tw,kw. (8)
172. ((alpha or beta or delta or gamma) adj3 variant? adj3 test*).tw,kw. (41)
173. (serologic* adj3 test*).tw,kw. (50287)
174. point of care testing/ (18292)
175. ((point-of-care or bedside? or bed side? or POC or rapid*) adj3 (assay? or immunoassay? or immuno-assay? or detect* or diagnos* or screen* or test*)).tw,kw. (226660)
176. (field adj3 (assay? or immunoassay? or immuno-assay? or detect* or diagnos* or screen* or test*)).tw,kw. (75079)
177. POCT.tw,kw. (5137)
178. (rapid adj3 (antigen* adj3 (assay? or immunoassay? or immuno-assay? or test*))).tw,kw. (3272)
179. (random* adj3 (assay? or immunoassay? or immuno-assay? or detect* or diagnos* or screen* or test*)).tw,kw. (88476)
180. random sample/ (13341)
181. sampling/ (79889)
182. (random* adj3 sampl*).tw,kw. (179585)
183. (pool* adj3 sampl*).tw,kw. (18959)
184. coronavirus disease 2019/ep [epidemiology] (25103)
185. coronavirus infection/ep [epidemiology] (23535)
186. incidence/ (742644)
187. prevalence/ (1098179)
188. (incidence or prevalen*).tw,kw. (3900074)
189. or/129-188 [SURVEILLANCE - BROAD] (9710465)
190. 128 and 189 (1666)
191. exp animal/ or exp animal experimentation/ or exp animal model/ or exp animal experiment/ or nonhuman/ or exp vertebrate/ (53905444)
192. exp human/ or exp human experimentation/ or exp human experiment/ (42359723)
193. 191 not 192 (11547438)
194. 190 not 193 [ANIMAL-ONLY REMOVED] (1263)
195. limit 194 to yr=”2020-current” (1090)
196. 195 use oemezd [EMBASE RECORDS] (451)
197. COVID-19/ (85559)
198. SARS-CoV-2/ (81242)
199. Coronavirus/ (13278)
200. Betacoronavirus/ (40986)
201. Coronavirus Infections/ (57260)
202. (COVID-19 or COVID19).ti,ab,kw. (258067)
203. ((coronavirus* or corona virus*) and (hubei or wuhan or beijing or shanghai)).ti,ab,kw. (9910)
204. (wuhan adj5 virus*).ti,ab,kw. (535)
205. (2019-nCoV or 19nCoV or 2019nCoV).ti,ab,kw. (3468)
206. (nCoV or n-CoV or “CoV 2” or CoV2).ti,ab,kw. (91681)
207. (SARS-CoV-2 or SARS-CoV2 or SARSCoV-2 or SARSCoV2 or SARS2 or SARS-2 or severe acute respiratory syndrome coronavirus 2).ti,ab,kw. (99672)
208. (2019-novel CoV or Sars-coronavirus2 or Sars-coronavirus-2 or SARS-like coronavirus* or ((novel or new or nouveau) adj2 (CoV or nCoV or covid or coronavirus* or corona virus or Pandemi*2)) or (coronavirus* and pneumonia)).ti,ab,kw. (36536)
209. (novel coronavirus* or novel corona virus* or novel CoV).ti,ab,kw. (18558)
210. ((coronavirus* or corona virus*) adj2 “2019”).ti,ab,kw. (58733)
211. ((coronavirus* or corona virus*) adj2 “19”).ti,ab,kw. (9089)
212. (”coronavirus 2” or “corona virus 2”).ti,ab,kw. (29846)
213. (OC43 or NL63 or 229E or HKU1 or HCoV* or Sars-coronavirus*).ti,ab,kw. (7730)
214. (coronavirus* or corona virus*).ti. (43970)
215. (”B.1.1.7” or “B.1.351” or “B.1.617” or “B.1.427” or “B.1.429”).ti,ab,kw. (627)
216. (”P.1” and (Brazil* or variant?)).ti,ab,kw. (3403)
217. ((alpha or beta or delta or gamma) adj3 variant?).ti,ab,kw. (11631)
218. or/197-217 [COVID-19] (330858)
219. vaccinated.ti,ab,kw. (97171)
220. inoculated.ti,ab,kw. (153204)
221. immuni#ed.ti,ab,kw. (119795)
222. post-vaccinat*.ti,ab,kw. (11858)
223. post-inoculat*.ti,ab,kw. (11624)
224. post-immuni*.ti,ab,kw. (3940)
225. ((after or already or full or fully or post or received) adj3 (immunis* or immuniz* or immunity or inoculat* or vaccin*)).ti,ab,kw. (185426)
226. (status* adj3 (immunis* or immuniz* or immunity or inoculat* or vaccin*)).ti,ab,kw. (17468)
227. or/219-226 [VACCINATED] (471883)
228. 218 and 227 (5617)
229. Health Surveys/ (249807)
230. ((health or population?) adj3 survey?).ti,ab,kw. (179817)
231. ((disease* or pandemic*) adj3 (monitor* or survey?)).ti,ab,kw. (56234)
232. ((COVID or COVID-19 or COVID19) adj3 (monitor* or survey?)).ti,ab,kw. (2268)
233. ((coronavirus* or corona virus*) adj3 (monitor* or survey?)).ti,ab,kw. (283)
234. ((2019-nCoV or nCoV or n-CoV or SARS-CoV-2 or SARS-CoV2 or SARSCoV-2 or SARSCoV2 or SARS2) adj3 (monitor* or survey?)).ti,ab,kw. (523)
235. ((BNT162 or BNT162-01 or BNT162a1 or BNT162b1 or BNT162b2 or BNT162c2) adj3 (monitor* or survey?)).ti,ab,kw. (4)
236. ((alpha or beta or delta or gamma) adj3 variant? adj3 (monitor* or survey*)).ti,ab,kw. (6)
237. *Epidemiological Methods/ (6485)
238. Epidemiological Monitoring/ (10263)
239. (epidemiolog* adj3 monitor*).ti,ab,kw. (3326)
240. Seroepidemiologic Studies/ (26333)
241. ((seroepidemiol* or sero-epidemiol*) adj3 (monitor* or survey* or study or studies)).ti,ab,kw. (7220)
242. (seromonitor* or sero-monitor* or serological monitor*).ti,ab,kw. (574)
243. (seroprevalen* or sero-prevalen* or serological prevalen*).ti,ab,kw. (46880)
244. (serosurveillan* or sero-surveillan*).ti,ab,kw. (1338)
245. (serosurvey? or sero-survey? or serological survey?).ti,ab,kw. (9021)
246. Wastewater-Based Epidemiological Monitoring/ (2419)
247. Waste Water/ (50317)
248. (sewage* or wastewater or waste water).ti,ab,kw. (182349)
249. Norman score?.ti,ab,kw. (81)
250. Data Collection/ (305772)
251. ((collect* or monitor*) adj3 data).ti,ab,kw. (1015431)
252. Public Health Practice/ (70070)
253. ((community health or public health) adj3 (practice? or activit* or endeavo?r?)).ti,ab,kw. (15865)
254. exp Population Surveillance/ (306931)
255. surveillance*.ti,ab,kw. (477093)
256. (biosurveillance* or bio-surveillance*).ti,ab,kw. (735)
257. Mass Screening/ (167946)
258. screening.ti,ab,kw. (1436256)
259. ((mass or population*) adj3 screen*).ti,ab,kw. (58593)
260. (screen* or detect* or identif* or recogni*).ti. (2128877)
261. ((early or earlier or earliest or ongoing or regular*) adj5 (screen* or detect* or identif* or recogni*)).ti,ab,kw. (526276)
262. (case finding? or casefinding?).ti,ab,kw. (13278)
263. Metagenomics/ (24172)
264. (ecogenomic* or eco-genomic* or metagenomic* or meta-genomic*).ti,ab,kw. (32988)
265. ((communit* or ecologic* or environment* or population) adj3 genomic*).ti,ab,kw. (11715)
266. COVID-19 Testing/ (5913)
267. ((COVID or COVID-19 or COVID19) adj3 test*).ti,ab,kw. (7783)
268. ((coronavirus* or corona virus*) adj3 test*).ti,ab,kw. (1061)
269. ((2019-nCoV or nCoV or n-CoV or SARS-CoV-2 or SARS-CoV2 or SARSCoV-2 or SARSCoV2 or SARS2) adj3 test*).ti,ab,kw. (7733)
270. ((BNT162 or BNT162-01 or BNT162a1 or BNT162b1 or BNT162b2 or BNT162c2) adj3 test*).ti,ab,kw. (8)
271. ((alpha or beta or delta or gamma) adj3 variant? adj3 test*).ti,ab,kw. (41)
272. (serologic* adj3 test*).ti,ab,kw. (50216)
273. Point-of-Care Testing/ (18292)
274. ((point-of-care or bedside? or bed side? or POC or rapid*) adj3 (assay? or immunoassay? or immuno-assay? or detect* or diagnos* or screen* or test*)).ti,ab,kw. (226456)
275. (field adj3 (assay? or immunoassay? or immuno-assay? or detect* or diagnos* or screen* or test*)).ti,ab,kw. (74971)
276. POCT.ti,ab,kw. (5130)
277. (rapid adj3 (antigen* adj3 (assay? or immunoassay? or immuno-assay? or test*))).ti,ab,kw. (3261)
278. (random* adj3 (assay? or immunoassay? or immuno-assay? or detect* or diagnos* or screen* or test*)).ti,ab,kw. (87480)
279. (random* adj3 sampl*).ti,ab,kw. (178562)
280. (pool* adj3 sampl*).ti,ab,kw. (18914)
281. COVID-19/ep [epidemiology] (14412)
282. Coronavirus Infections/ep [epidemiology] (23494)
283. Incidence/ (742644)
284. Prevalence/ (1098179)
285. (incidence or prevalen*).ti,ab,kw. (3893916)
286. or/229-285 [SURVEILLANCE - BROAD] (9644667)
287. 228 and 286 (1610)
288. exp Animals/ not Humans/ (16106641)
289. 287 not 288 [ANIMAL-ONLY REMOVED] (1204)
290. limit 289 to yr=”2020-current” (1050)
291. 290 use coch [CDSR RECORDS] (0)
292. 290 use cctr [CENTRAL] (78)
293. 96 or 196 or 291 or 292 [ALL DATABASES] (1086)
294. remove duplicates from 293 (737) [TOTAL UNIQUE RECORDS]
295. 294 use ppez [MEDLINE UNIQUE RECORDS] (541)
296. 294 use oemezd [EMBASE UNIQUE RECORDS] (126)
297. 294 use coch [CDSR UNIQUE RECORDS] (0)
298. 294 use cctr [CENTRAL UNIQUE RECORDS] (70)

### Web of Science

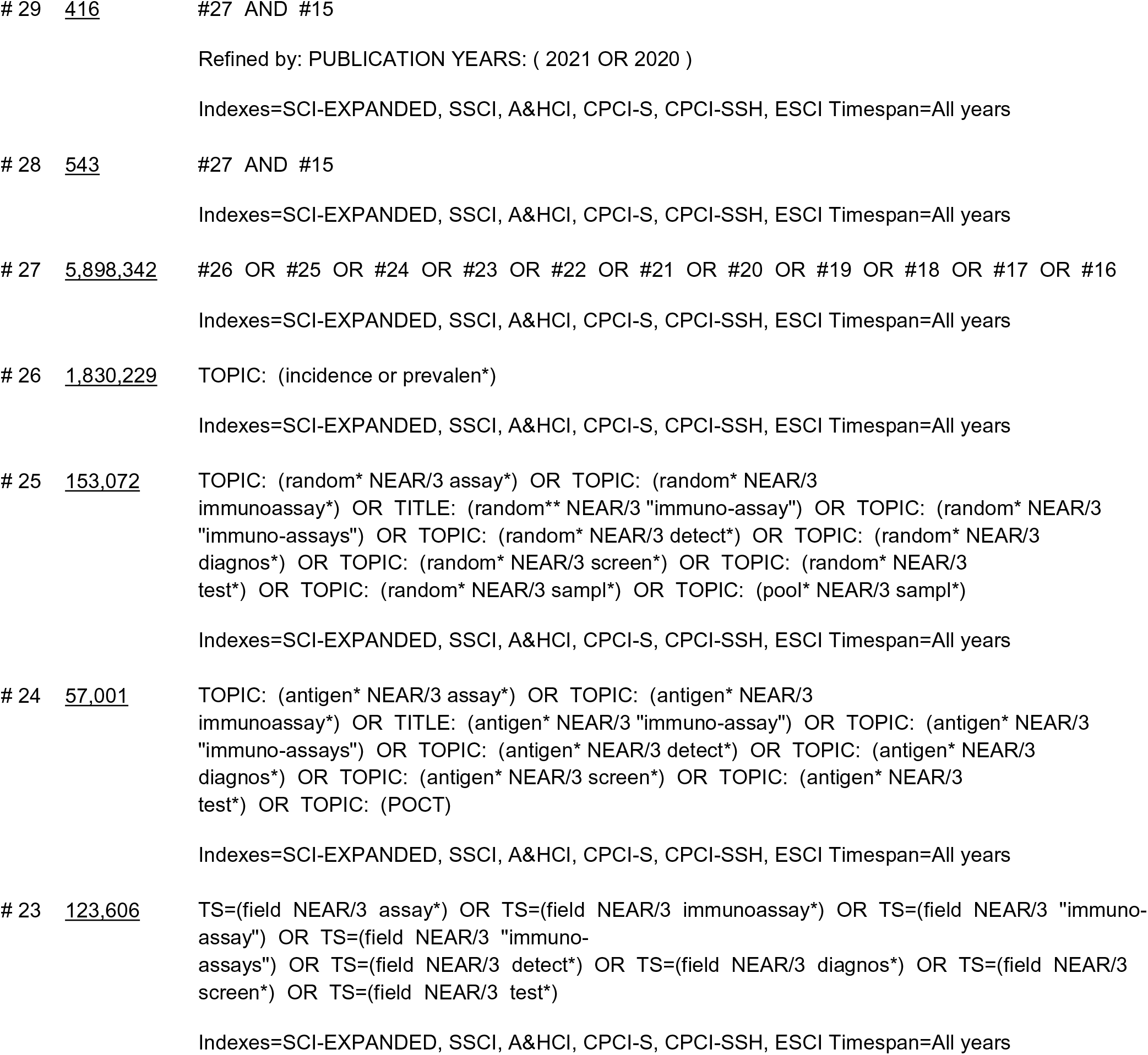
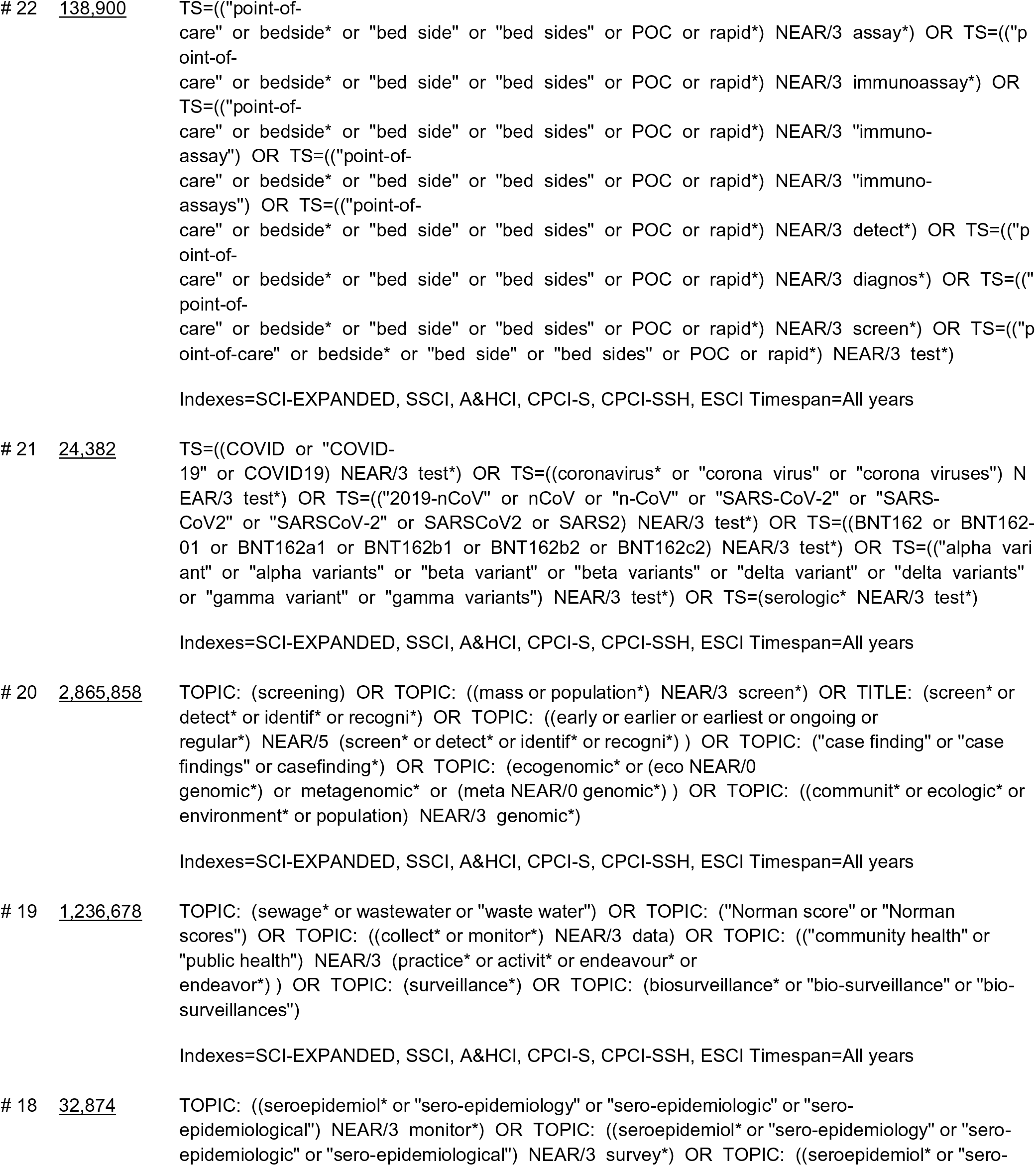
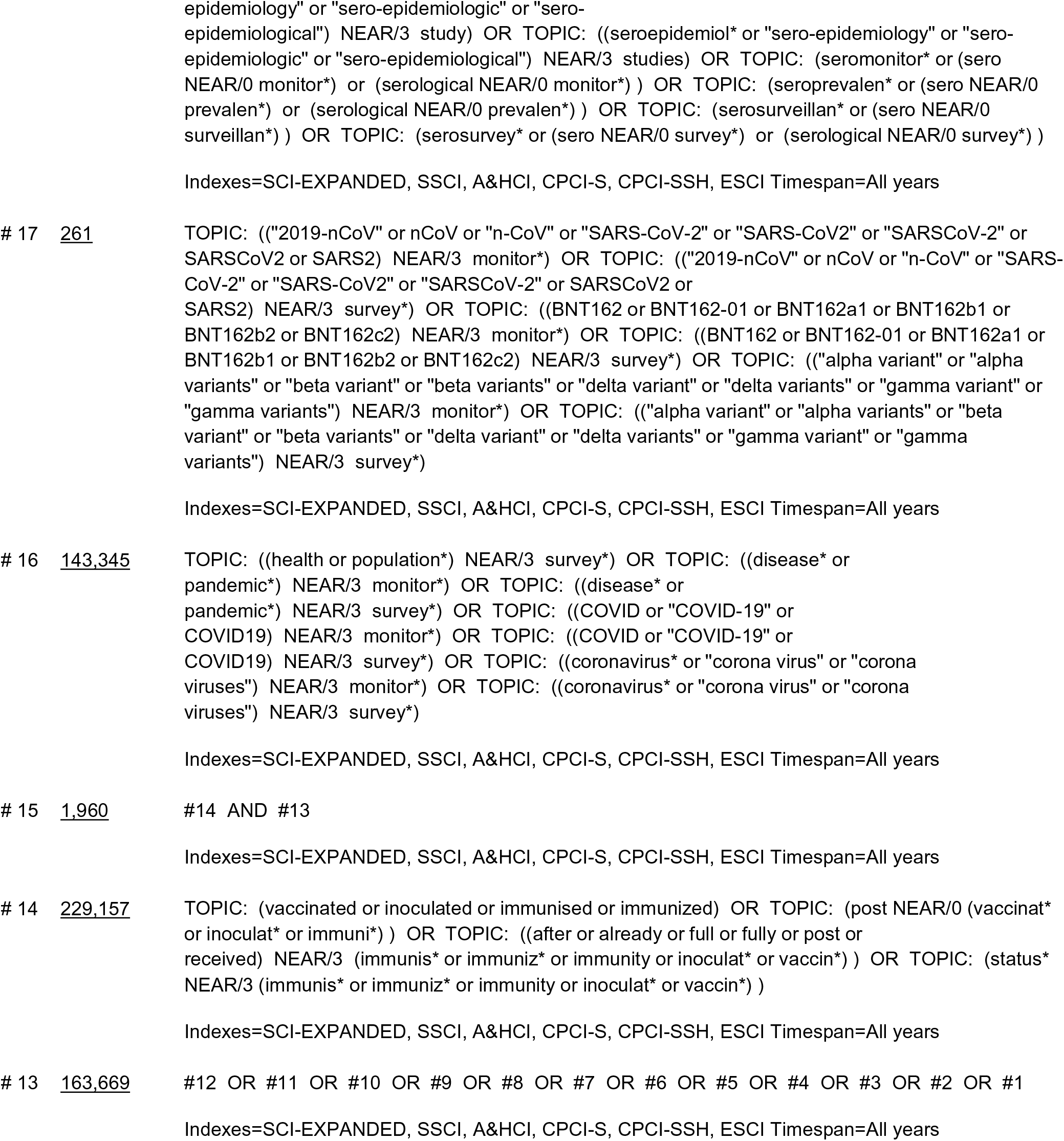
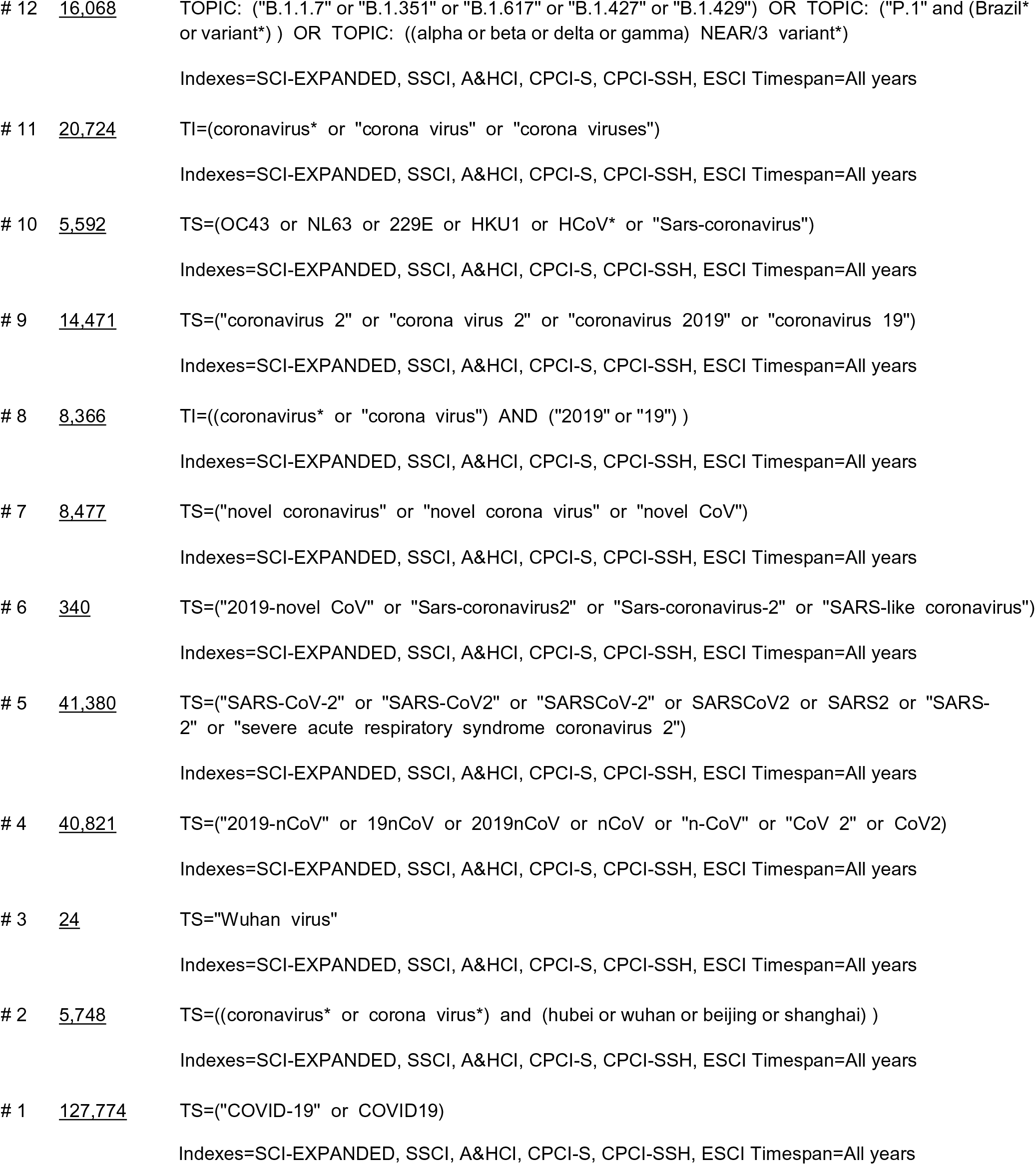

## Appendix B: Additional Details on International Guidance

**Table 6:** Summary of Included Guidance

| Country | Institute/Author | Scope, Setting, Evidence | Surveillance Method Used | Summary of Guidance |
| --- | --- | --- | --- | --- |
| June 2021 |  |  |  |  |
| Australia (Southern) | Government of South Australia - SA Health<sup>52</sup> | Scope: Regional<br>Setting: Community<br>Level of Evidence: Unclear | Wastewater surveillance<br>**Stated reason for use:** Testing wastewater can help provide an early warning signal of COVID-19 infections in people living in a particular wastewater catchment area. | Wastewater testing is done to support the State's response to COVID-19; helps to provide an early warning signal of COVID-19 infections in people living in a particular wastewater catchment area. |
| Australia (Western) | Government of Western Australia - Department of Health<sup>53</sup> | Scope: Regional<br>Setting: Community<br>Level of Evidence: Unclear | Wastewater surveillance<br>**Stated reason for use:** Wastewater testing will complement – but cannot replace – other kinds of COVID-19 testing, including nose and throat swabs. Wastewater testing indicates if COVID-19 may be present in the broader community. | The wastewater testing program complements other work already being done to monitor COVID-19, including testing of individual people using nose and throat swabs, and, together, these help inform WA Health's response to the pandemic. Samples are taken from six metropolitan locations and 10 regional localities. |
| Germany | Federal Ministry of Health<sup>54</sup> | Scope: National<br>Setting: Community<br>Level of Evidence: Unclear | Digital support for health authorities, digital symptom diary, contact management<br>**Stated reason for use:** Enable nationwide, secure and fast electronic reporting and information processing with regard to positive SARS-CoV2. In this way, the effort for the reporting laboratories and the responsible health authorities and state authorities is to be reduced, and infection events that occur are to be contained by the possibility of introducing targeted infection protection measures more quickly. | German Electronic Reporting and Information System for Infection Protection (DEMIS) for reporting laboratory confirmed cases.<br>Digital symptom diary application (Climedo) which is used to conduct daily surveys of contacts of persons with COVID-19.<br>Surveillance Outbreak Response Management and Analysis System (SORMAS): For efficient identification of contacts. |
| May 2021 |  |  |  |  |
| UK | Public Health England<sup>55</sup> | Scope: National<br>Setting: School, healthcare setting, community<br>Level of Evidence: Evidence informed | Serosurveillance, population PCR<br>**Stated reason for use:** Children represent a small proportion of the total number of confirmed COVID-19 cases. There are limited data on childhood SARS-CoV-19infections, especially from Europe. Public Health England (PHE) along with NHS partners and academic collaborators has implemented a number of ongoing surveillance programmes to monitor the course, progression and outcomes ofCOVID-19 in children. | Surveillance methods include:<br>• SKIDs PLUS: COVID-19 surveillance in secondary schools (2020/21 academic year): Participants in 20 secondary schools will have nasal swabs, saliva and blood samples for virus and antibody testing at recruitment and end of term.<br>• Outbreak surveillance in educational settings (Ongoing): PHE has been monitoring cases, clusters and outbreaks in educational settings since the reopening of schools on 1 June 2020.<br>• School infection surveys: PHE conducting a large prospective school survey of staff and students in up to 150 schools across England.<br>• Electronic notification of Cases: PHE receives electronic notifications of all confirmed COVID-19 cases from NHS hospital laboratories in England through the Second-Generation Surveillance System (SGSS).<br>• Clinical follow up of laboratory-confirmed cases in neonates up to 28 days of age: Pediatricians across the UK and Ireland will receive weekly emails from the British Paediatric Surveillance Unit (BPSU) team to report whether they have managed a case of neonatal COVID-19 in the previous week. Those confirming a case will be asked to complete a short clinical questionnaire.<br>• RAPID-19 Study (COVID in children of HCWs): PHE is working with teams across the UK to conduct surveillance in children of healthcare workers across 5 cities. Samples will be taken for SARS-CoV-2 antibodies at recruitment, around 8 weeks later and 4 to 6 months later. This will allow us to monitor how much<br>the children have been exposed to the virus between the different testing periods. |
| USA | CDC<sup>56</sup> | Scope: National<br>Setting: Community<br>Level of Evidence: Evidence informed | Population genomic screening<br>**Stated reason for use:** Improve our understanding of which variants are circulating the US, how quickly variants emerge, and which variants are the most important to characterize and track in the terms of health. | CDC contracted large commercial diagnostic labs to sequence samples across the US to sequence SARS-CoV-2 genomes. Goal is to identify and characterize variant viruses (either new ones identified in the US or those already identified abroad) and to investigate how variants impact COVID-19 disease severity and the effectiveness of vaccines, treatment, and diagnostic tests. |
| April 2021 |  |  |  |  |
| Australia | Communicable Diseases Network Australia<sup>57</sup> | Scope: National<br>Setting: Community<br>Level of Evidence: Evidence informed | Case-based reporting, surveillance of outbreaks and clusters, targeted active case finding, syndromic and sentinel surveillance, serosurveillance, virus genomics, monitoring of personal behaviours<br>**State reason for use:** Australia continues to follow a suppression strategy in response to COVID-19. Each disease surveillance goal contributes information, via key indicators, that support strategic and operational decision-making by national and jurisdictional governments and public health authorities. | Surveillance strategies include:<br>Monitoring the characteristics and time trends of COVID-19 cases to support planning and evaluation of prevention activities and testing services.<br>Providing updates on the SARS-CoV-2 transmission potential and forecasts of epidemic activity to support planning and evaluation of prevention activities.<br>Characterizing clusters and secondary cases to facilitate management within and across jurisdictions and inform targeted control strategies.<br>Monitoring testing counts for SARS-CoV-2 and to provide denominator to track how effectively testing is being utilized and for an indication of the positivity yield.<br>Monitoring community trends in "fever/acute respiratory illness (ARI)" and the proportion tested and attributable to SARS-CoV-2 to assess the extent of community transmission and the effectiveness of public health measures aimed at both prevention and case detection.<br>Determining seroprevalence rates of SARS-CoV-2 by population group and geographic location to<br>complement our understanding of population transmission of SARS-CoV-2 from other surveillance data.<br>• Reporting on strategically targeted asymptomatic testing of selected populations potentially at higher risk of exposure.<br>• Monitoring the characteristics of deaths due to COVID-19 as a key measure of the health impact of the disease in our community.<br>• Describing the clinical severity of COVID-19 cases to allow prediction of health resource use and to standardize models of care, and characterizing risk factors for serious infection to allow for targeted public health measures.<br>• Assessing case detection and contact tracing to understand capacity to detect and interrupt chains of transmission and to support resource planning.<br>• Monitoring adherence to recommended behavioural practices to support planning and evaluation of prevention activities.<br>• Measuring impacts on the tertiary health care system to contribute to forecasts of capacity requirements to redirect resources as required. |
| France | Sante Publique France<sup>58</sup> | Scope: National, outside of Brittany<br>Setting: Community<br>Level of Evidence: Unclear | Population PCR screening, with sequencing of SA variant cases<br>**Stated reason for use:** to describe probable and confirmed cases linked to the B.1.616 variant and to monitor the geographical expansion of this variant outside of Brittany to adapt appropriate measures. | • Physicians to report suspected B.1.616 cases to ARS (Regional health authority); investigation into these cases to be conducted by ARS and regional units of Public Health France; cases that are negative for nasopharyngeal swab but positive for deep swab are to be sent for sequencing. |
| India | Ministry of Health and Family Welfare, Government of India<sup>59</sup> | Scope: National<br>Setting: Community<br>Level of Evidence: Unclear | Population genomic screening<br>**Stated reason for use:** Gather whole genome sequencing information in the community by targeting events like clustering of COVID cases, suspected super-spreader events, clustering of cases in institutions, suspected vaccine failure and re-infection clusters etc. | The objective of the special surveillance is to gather WGS information in the community by targeting events like clustering of covid cases, suspected super-spreader events, clustering of cases in institutions, suspected vaccine failure and re-infection clusters etc.<br>Each site to send 15 random samples every 15 days to designated lab for WGS. The samples from the tertiary care health facilities should include samples of patients with severe illness/prolonged admission, suspected re-infection, suspected vaccination failure/infection following vaccination, any other special case etc. |
| International | Pan American Health Association/WHO<sup>60</sup> | Scope: International<br>Setting: Community<br>Level of Evidence: Evidence informed | Population PCR screening for both influenza and COVID-19, population genomic screening<br>**Stated reason for use:** The threat of influenza epidemics and pandemics persists. It is imperative for the WHO's Global Influenza Surveillance and Response System to maintain meaningful surveillance of influenza worldwide and for countries to remain vigilant while adapting to meet COVID-19 surveillance objectives. | Operational considerations for surveillance of severe acute respiratory illness, and/or influenza-like illnesses using multiplex assay, including prioritization of test use; operational considerations for laboratories including clinical sample collection, storage, and transport; Multiplex assay approach including how to interpret and report results; Guidelines on testing algorithms; Influenza and COVID-19 surveillance data reporting including frequency of reporting (e.g., weekly for COVID-19 testing data) and where data should be submitted. |
| USA | CDC<sup>61</sup> | Scope: National<br>Setting: Community<br>Level of Evidence: Unclear | Population genomic screening<br>**Stated reason for use:** National and global sequencing efforts have identified changes in the SARS-CoV-2 genetic code resulting from transmission and evolution in humans and animals. These changes can affect many aspects of our response including transmission, diagnostics, therapeutics, and vaccines. | CDC guidelines for jurisdictions to send in positive PCR samples to conduct further genomic testing, including how specimen should be collected, stored, and transported. |
| March 2021 |  |  |  |  |
| France | Sante Publique France<sup>62</sup> | Scope: National<br>Setting: Healthcare<br>Level of Evidence: Unclear | Population PCR screening, with sequencing of SA variant cases<br>**Stated reason for use:** To track the epidemiologic characteristics of severe influenza and COVID-19 cases admitted to the ICU; to document the specific contribution of influenza and SARS-CoV-2 viruses among ICU admissions; and to describe and document the comorbidities and characteristics of patients admitted to the ICU for influenza or SARS-CoV-2 infection. | Surveillance of severe COVID cases; All COVID cases admitted to ICU captured in SI-VIC; linked to other datasets to obtain more identifying information and track epidemiologic characteristics of the cases, describe comorbidities, and document extent of COVID contribution to ICU admissions; Centre national de référence (CNR) to sequence the cases.<br>To be carried out by network of sentinel ICUs established in 2018; sentinel ICU to report case to regional unit of Public Health France who will conduct follow-up. |
| UK | Public Health England<sup>63</sup> | Scope: National<br>Setting: Community, health care setting, long-term care<br>Level of Evidence: Evidence informed | PCR screening, serosurveillance, hospital screening, genome sequencing<br>**Stated reason for use:** To monitor coverage of the vaccine in targeted populations and identify under vaccinated groups; to rapidly detect and evaluate possible adverse events associated with vaccination; to estimate the effectiveness of the vaccine at preventing a spectrum of disease outcomes and onwards transmission in different targeted populations, and against different viral variants, as well as the duration of any protective effect; to identify risk factors for and outcomes of vaccine failure, including any impact on strain evolution; to monitor the overall impact of the vaccination programme on COVID-19 in the wider population including the indirect effect on groups not targeted by the vaccination programme; to monitor the impact of the vaccination programme on prevalence of antibodies against COVID-19 as an indicator of population level immunity, and to monitor antibody waning in the population; to monitor attitudes to vaccination and identify barriers to high vaccine uptake, and; to monitor inequalities in each of these outcome measures | Surveillance strategies include:<br>Routine COVID-19 testing through the Second-Generation Surveillance System (SGSS) linked to vaccination data from the NIMS to provide a dataset for monitoring vaccine effectiveness using a test-negative case control approach by vaccine, age group, clinical co-morbidities and different dosing schedules that may be used in the program.<br>Enhanced surveillance: Once a vaccination program is implemented and the earliest eligible groups have been offered a full course of vaccination, PHE will begin enhanced surveillance of a subset of cases in vaccine eligible groups identified through the routine testing. Clinical questionnaires will be completed with the case and their GP or hospital clinician on vaccination history, past medical history, symptoms and outcomes. Repeat nose and throat swabs and acute and convalescent serum and oral fluid samples will be taken, these will be used to confirm recent infection, provide evidence of immune response following vaccination and identify<br>primary and secondary vaccine failures, estimate viral load, viral replication and culturable virus as markers of infectiousness, test for other respiratory viruses, and undertake sequencing to identify nucleotide changes that may favour vaccine escape and differences in phenotype.<br>• GP electronic health record studies: In addition to monitoring clinical presentations with respiratory syndromes, the network undertake sentinel swabbing of patients presenting with influenza like illness.<br>• Hospitalization and mortality rates will be monitored as surveillance of vaccine effectiveness against severe disease.<br>• Surveillance studies: number of studies have been established since the start of the pandemic with routine asymptomatic infection and these will be used to monitor vaccine effectiveness against infection: SIREN, Vivaldi, Community infection survey, and routine data sources.<br>• Transmissibility will be monitored through analysis of CT values, culturable virus and duration of PCR positivity in vaccinated cases; to directly monitor the effect on transmission, a sample of cases identified through enhanced surveillance as well as healthcare workers and care home staff identified through the SIREN (Sarscov2 Immunity & REinfection EvaluationN) study and the Vivaldi study (described below) will be recruited to monitor the effect of vaccination on their risk of onwards transmission. Routine testing data will also be used to monitor the number of secondary cases in households where the first case identified in the<br>household is vaccinated using the PHE HOSTED dataset with linkage to the NIMS data.<br>• Protocols for monitoring vaccine effectiveness in outbreaks in closed settings have been developed.<br>• Other surveillance methods include surveillance of vaccine failure (including follow-up with viral whole genome sequencing) and sero-surveillance (including blood donor samples, routine blood tests, and residual sera). |
| February 2021 |  |  |  |  |
| International | Pan American Health Association/WHO<sup>64</sup> | Scope: International<br>Setting: Community<br>Level of Evidence: Evidence informed | Population genomic screening<br>**Stated reason for use:** A time series of samples is necessary to detect trends of change in genetic diversity and emerging variants. As well, appropriate number and representative samples, including unusual case samples, are important for generating reliable genomic sequencing data. | • Guidelines and criteria for sampling representative data, virologic characteristics of clinical samples, timeframe and number of SARS-CoV-2 samples. When doing genomic sequencing of samples, should consider a submission of ≥50 samples per month from varying demographic groups. |
| Spain | Ministry of Health – Spain<sup>65</sup> | Scope: National<br>Setting: Community, health care settings, long term care, prisons<br>Level of Evidence: Evidence informed | PCR screening, rapid antigen screening, genomic screening<br>**Stated reason for use:** Early detection of cases with active SARS-CoV-2 infection; the early establishment of necessary control measures to prevent new infections; the availability of the necessary information for epidemiological surveillance, with a level of adequate disaggregation and detail. | • Genomic sequencing: to establish a surveillance of the different phylogenetic variants of COVID, its distribution, transmission and public health implications, a protocol the integrates genomic sequencing into the COVID surveillance system has been developed to detect variants.<br>• Spot screenings: targeting specific groups or populations via PCR nasopharyngeal swabs.<br>• Periodic screening: of workers and residents of health or social health centres using PCR.<br>• Systematic screening: for new admissions in health or social health centres or prior to care actions. |
| USA | CDC<sup>66</sup> | Scope: National | Serosurveillance | • Using seroprevalence surveys to learn about the total number of people that have been infected, including |
|  |  | Setting: Community<br>Level of Evidence:<br>Evidence informed | **Stated reason for use:** To better understanding how many infections with SARS-CoV-2 have occurred at different time points, in different locations, and within different populations in the US. | those infections that may have been missed. These surveys can also help estimate how much of the population has not yet been infected, helping public health officials plan for future healthcare needs. |
| January 2021 |  |  |  |  |
| Europe | ECDC<sup>67</sup> | Scope: International<br>Setting: Community<br>Level of Evidence:<br>Evidence informed | Genomic surveillance (Pre-screening RT-PCR before testing for emerging variants)<br>**Stated reason for use:** To detect introduction of known variants and control the spread and impact of emerging variants. | • Systematic sequencing of a representative or random selection of detected viruses, which should be coordinated regionally and nationally. Laboratories should consider implementing pre-screening RT-PCR approaches to detect N501Y or S-gene target failure (deletion 69-70) variant viruses.<br>• All laboratories should be requested to report their results to the national public health institute that coordinates the collection of information. National public health authorities should notify cases of the variants of concern through the Early Warning and Response System (EWRS), and TESSy for case-based surveillance and aggregate reporting (which has been adapted for this purpose).<br>• To be able to confirm infection with a specific variant, sequencing of the whole SARS-CoV-2 genome, or at least the S-gene for the current variants, is required.<br>• For the VOC 202012/01, a negative S-gene result in multiplex RT-PCR assays, with positive results for the other targets, has been used as an indicator or pre-screening method to identify the variant. However, it should be noted that this target failure is not exclusive to VOC 202012/01, and confirmation using sequencing is always recommended.<br>• Increasing the numbers of sequenced samples pre-screened by S-gene target failure can be considered to<br>assess the regional correlation between S-gene target failure and VOC 202012/01 as this varies with the regionally circulating variants. If the correlation is very high, S-gene target failure can be used to approximate the frequency of VOC 202012/01.<br>• A minimum ability to roughly quantify the proportion of a variant present at a prevalence of 2.5% of the total circulating variants is recommended. This requires each country to sequence at least around 500 randomly selected samples each week.<br>• Sequencing of samples from large outbreaks and samples connected to travelers (either from point of entry screening or outbreaks involving a traveler) should be prioritized.<br>• Sequencing should be prioritized for individuals who present with a 'breakthrough infection' identified >14 days after receiving the first dose of COVID-19 vaccine (see section on Vaccination).<br>• Viral isolation of variants of SARS-CoV-2 should be carried out in P3 (Biosafety level (BSL) laboratories to prevent the accidental dissemination of a variant through laboratory exposure.<br>• In general, laboratory preparedness should be among the current high priorities, and laboratories should: 1) consider implementing diagnostic pre-screening for variants of concern (e.g. N501Y and deletion 69-70); 2) ensure human and material resources are available to manage an increasing number of requests for detection and characterization of SARS-CoV-2 samples; 3) increase sequencing capacity by making use of all possible sequencing capacity in the Member States<br>from clinical, diagnostic, academic and commercial laboratories, or requesting assistance from ECDC. |
| France | Sante Publique France<sup>68</sup> | Scope: National<br>Setting: Community, points of entry for travel<br>Level of Evidence: Unclear | Population PCR screening, with sequencing of UK and SA variant cases<br>**Stated reason for use:** To document the circulation of the UK variant and the South African variant in France and slow their spread. | Daily monitoring of cases in individuals coming from the UK or South Africa entered into SI-DEP to identify cases coming from abroad.<br>Any biologic material from a new probable UK or SA variant case with a CT value <25 or a confirmed case must be immediately sent to Centre National de Reference (CNR; Laboratoire de l'Institut Pasteur or Laboratoire de Lyon) for sequencing. Confirmed cases are sent to Public Health France and Centre de crise sanitaire (CCS)<br>Surveillance geared towards variant infections in young adults, including closely monitoring infections in <30-year-olds, collecting samples in educational institutions (including PCR and antigen tests), sending positive case samples for sequencing, etc |
| Spain | Ministry of Health – Spain<sup>69</sup> | Scope: National<br>Setting: Community<br>Level of Evidence: Evidence informed | Population genomic screening<br>**Stated reason for use:** Determine the incidence of variants of interest for public health; early identification of new variants that present increased transmissibility/virulence, vaccine breakthrough, or phenotypic change; identification of vulnerable groups associated with new variants; carry out viral phylodynamic studies. | For the identification and monitoring of the circulating variants in the country, a procedure is proposed based on the principles: complete sequencing shall be carried out and done in a planned manner, including a representative number of cases from all AACs and a laboratory will be established.<br>In addition to the sequencing of COVID in a representative number of cases, it is important to detect early new variants of public interests and therefore sequencing of the following cases/situations will be<br>indicated: suspected reinfection, suspicion of new variants of interest to public health infection, cases with suspected infection with variants that escape immunity, situations where high transmissibility or virulence is suspected.<br>Samples used for genetic testing come from nasopharyngeal swabs. |
| December 2020 |  |  |  |  |
| International | WHO<sup>70</sup> | Scope: International<br>Setting: Community, health care settings, sentinel sites, closed settings, travelers at points of entry<br>Level of Evidence: Evidence informed | Nucleic acid amplification tests, antigen-detecting rapid diagnostic tests, Antibody detection (serology), reporting/epidemiological data<br>**Stated reason for use:** The aim of national surveillance for COVID-19 is to enable public health authorities to reduce transmission of SARS-CoV-2, thereby limiting associated morbidity and mortality. The objectives of COVID-19 surveillance are to: Enable rapid detection, isolation, testing, and management of cases; detect and contain clusters and outbreaks, especially among vulnerable populations; identify, follow-up and quarantine contacts; guide the implementation and adjustment of targeted control measures, while enabling safe resumption of economic and social activities; evaluate the impact of the pandemic on health care systems and society; monitor longer term epidemiologic trends and evolution of SARS-CoV-2 virus and monitor trends in covid-19 deaths, and; contribute to the understanding of the co-circulation of SARS-CoV-2 virus, influenza and other respiratory viruses, and other pathogens. | Surveillance in the community: systematic detection and reporting of events of public health significance within a community by community members. This can be through participation in contact tracing and cluster investigations.<br>Surveillance at the primary care level: where possible, testing should be available at primary care clinics or complementary options should be available to establish dedicated SARS-CoV-2 community testing facilities, such as drive-through sites or fixed sites in community buildings.<br>Hospital based surveillance: patients with probable or confirmed COVID-19 admitted to hospitals should be notified to national public health authorities within 24 hours of identification.<br>Sentinel site surveillance: syndromic surveillance and collection of respiratory specimens using existing case definitions through sentinel networks. Laboratories should continue virologic testing of routine sentinel site samples for influenza, with the addition of testing samples for SARS-CoV-2. Countries are encouraged to conduct year-round sentinel surveillance for acute respiratory syndromes with testing of samples for |
|  |  |  |  | SARS-CoV-2. Patients selected for additional testing should preferably be representative of the population and include all ages and both sexes.<br>• Closed settings: high risk groups residing or working in closed settings is necessary to ensure the prompt detection of cases and clusters faster than through primary-care or hospital-based surveillance (prisons, residential facilities, retirement communities and care homes for persons with disabilities).<br>• Health care-associated infections: All cases and clusters in health care settings should be investigated and documented for their source and transmission patterns to allow rapid control.<br>• Mortality surveillance: number of COVID deaths occurring in hospitals should be reported daily.<br>• Laboratory surveillance: Nucleic acid amplification tests, Antigen detecting rapid diagnostic tests, antibody detection (serology).<br>• Participatory surveillance: Participatory disease surveillance enables members of the public to self-report signs or symptoms, without laboratory testing or assessment by a health care provider. |
| **November 2020** |  |  |  |  |
| Europe | ECDC<sup>71</sup> | Scope: International<br>Setting: Community, healthcare setting<br>Level of Evidence: Evidence informed | Population rapid antigen screening<br>**Stated reason for use:** Rapid antigen tests can offer a significant advantage over RT-PCR in terms of bringing testing closer to persons to test and timeliness of results. Benefits of rapid antigen testing include: prompt clinical management of cases with COVID-19-compatible symptoms at admission; control transmission – early detection of cases, contact tracing, | • Considerations for the use of rapid antigen tests in settings of low and high infection prevalence and the need for confirmatory testing: In a high prevalence setting, rapid antigen tests will have a high PPV. In such a situation, a positive result from a rapid antigen test (even with a lower specificity than in RT-PCR tests and thus a higher probability of false positivity) is likely to indicate a true infection and may not require |
|  |  |  | population-wide testing; mitigate the impact of COVID-19 in healthcare and social-care settings: triage at admission, early detection and isolation; identify clusters or outbreaks in specific settings: early detection and isolation. | confirmation by RT-PCR. --On the other hand, any negative test result should be confirmed by RT-PCR immediately or, in case of unavailability of RT-PCR, with another rapid antigen test a few days later (to allow the viral load to increase in previously false negative result). This is particularly true for asymptomatic cases with a known history of exposure.<br>• In any high-risk settings with vulnerable populations only RT-PCR should be used, unless RT-PCR capacity is limited. In vulnerable populations with symptoms, multiplex RT-PCR would be best suited for confirmation to exclude symptoms caused by other respiratory pathogens.<br>• In a low prevalence setting, rapid antigen tests will have a high NPV but a low PPV. Therefore, if used correctly, rapid antigen tests should be able to rule out a highly infectious case in such a setting. A negative test result may not require confirmation by RT-PCR, whereas a positive test will need immediate sampling for a confirmation by RT-PCR.<br>• Recurring testing by rapid antigen test every 2-3 days with the aim to identify infectious cases in a population can partly mitigate the lower sensitivity of the test and can be used in certain settings such as in staff of health care settings.<br>• In low prevalence settings, sufficient RT-PCR and logistics capacity will probably be in place to ensure a rapid turnaround of results. However, there could still be an added value to the use of rapid antigen tests because of the low cost and rapid turnaround time of analysis. Here, a careful cost-benefit calculation has to<br>be made in order not to exhaust the overall testing capacity in settings which have low impact on the course of the epidemic and the resources should rather be reserved for settings where highly infectious persons need to be detected.<br>• Testing in symptomatic patients: detailed guidance is provided in the document.<br>• Testing in asymptomatic patients: detailed guidance is provided in the document.<br>• Rapid antigen tests can be used for screening and serial testing (every two to three days) of residents and staff in healthcare, home care, long-term care facilities, closed settings (e.g. prisons, migrant detention and reception centres) and occupational settings in areas in which there is ongoing community transmission. Additional guidance is provided in the document. |
| UK | Wade et al. Summary for SAGE<sup>72</sup> | Scope: National<br>Setting: Community<br>Level of Evidence: Evidence informed | Wastewater surveillance<br>**Stated reason for use:** Wastewater surveillance is a reliable, timely and cost-effective method to serve the needs of public health. | • The English national surveillance programme samples from 44 STW, with data available since 22/07/2020, sampling a frequency of four measurements per week. Across all programmes, viral RNA data in the wastewater is being used along with complementary data, such as catchment area, sewer hydrology and sample inorganics such as ammonia and orthophosphate, to explore the relationship between SARS-CoV-2 RNA levels in STW influent and clinical cases in the population. |
| USA | CDC<sup>73,74</sup> | Scope: National<br>Setting: Community | Wastewater surveillance<br>**Stated reason for use:** To complement other COVID-19 surveillance indicators that inform public health actions. | • Wastewater surveillance is used to complement other COVID-19 surveillance indicators to inform public health actions. No interventions or public health actions should be solely based on wastewater data. |
|  |  | Level of Evidence:<br>Evidence informed |  |  |
| **August 2020** |  |  |  |  |
| Brazil | Ministry of Health Brazile (Ministerio de Saude) <sup>75</sup> | Scope: National<br>Setting: Community<br>Level of Evidence:<br>Unclear | Case-based reporting and laboratory reporting of positive cases;<br>Sentinel surveillance via Influenza sentinel surveillance network<br>**Stated reason for use:** Early identification of the occurrence of COVID-19 cases; establish criteria for the notification and registration of suspected cases in health services, public and private; establish procedures for laboratory investigation; monitor and describe the pattern of morbidity and mortality from COVID-19; monitor the clinical and epidemiological characteristics of the SARS-CoV-2 virus; establish prevention and control measures, and; carry out timely and transparent communication of the epidemiological situation in Brazil | Surveillance objectives include:<br>• Early identification of the occurrence of COVID-19 cases<br>• Establish criteria for the notification and registration of suspected cases in health services, public and private<br>• Establish procedures for laboratory investigation<br>• Monitor and describe the patter of morbidity and mortality from COVID-19<br>• Monitor the clinical and epidemiological characteristics of the SARS-CoV-2 virus<br>• Establish prevention and control measures<br>• Carry out timely and transparent communication of the epidemiological situation in Brazil |
| USA | CDC<sup>76</sup> | Scope: National<br>Setting: Community<br>Level of Evidence:<br>Evidence informed | Hospitalization secondary to COVID-19<br>**Stated reason for use:** Coronavirus Disease 2019 Associated Hospitalization Surveillance Network (COVID-NET) is a population-based surveillance system that collects data on laboratory-confirmed COVID-19-associated hospitalizations among children and adults through a network of over 250 acute-care hospitals in 14 states. COVID-NET is CDC's source for important data, and provides important clinical information on COVID-19-associated hospitalizations, including age group, sex, race/ethnicity and underlying health conditions. | • COVID-NET is a population-based surveillance system meant to collect, analyze, and interpret data regarding lab-confirmed Covid-19-associated hospitalizations among children and adults. Cases are identified in COVID-Net if they test positive through a test ordered by a health care professional and are hospitalized within 14 days of the positive test. Clinical data collected using a standardized case reporting form by trained surveillance officers, so the data are in a standardized and uniform way. COVID-Net comprises of 99 counties in 14 states (10% of the population). |
| **April 2020** |  |  |  |  |
| Europe | ECDC<sup>77</sup> | Scope: International | Symptoms and population PCR screening | • In countries comprehensively testing suspected cases for COVID-19, the most accurate indicators of intensity |
|  |  | Setting: Community, healthcare settings, long-term care<br>Level of Evidence: Evidence informed | **Stated reason for use:** Monitor the intensity, geographic spread and severity of COVID-19 in the population in order to estimate the burden of disease, assess the direction of recent time trends, and inform appropriate mitigation measures; monitor viral changes to inform drug and vaccine development, and to identify markers of severe infection; monitor changes in which risk groups are most affected in order to better target prevention efforts; monitor the epidemic's impact on the healthcare system to predict the trajectory of the epidemic curve and inform resource allocation and mobilization of surge capacity as well as external emergency support; monitor the impact of any mitigation measures to inform authorities so they can adjust the choice of measures, as well as their timing and intensity; detect and contain nosocomial outbreaks to protect healthcare workers and patients, and; detect and contain outbreaks in long-term care facilities and other closed communities to protect those most at risk of severe disease and poor outcomes. | will be the absolute number of newly confirmed cases and their notification rate per 100 000 population.<br>• Sentinel syndromic surveillance: should integrate COVID-19 surveillance with sentinel surveillance of influenza-like illness (ILI) or acute respiratory infection (ARI), which is in place in most EU/EEA Member States. The naso-pharyngeal swabs obtained by sentinel physicians from a systematic sample of patients presenting with ILI/ARI should also be tested for severe acute respiratory syndrome coronavirus 2 (SARS-CoV-2) in addition to influenza virus and other respiratory viruses. In countries where sentinel physicians are not able to swab their patients, other approaches can be considered, such as self-swabbing and shipping of specimens using dedicated channels.<br>• Helplines, surveys, participatory surveillance. Countries not systematically testing most suspected cases while limiting physical access to primary healthcare (for example by encouraging people to call specific COVID-19 helplines, or when people are placed in lockdown) should consider analysing data from alternative sources. These could include phone consultations of sentinel physicians, calls to regional/national healthcare telephone helplines, consultations of online healthcare apps or self-assessment tools for advice on COVID-19 testing, or population-based participatory syndromic surveillance schemes for influenza that exist in a number of Member States. Resources permitting, countries can also conduct their own regular telephone surveys. |
|  |  |  |  | • Virological sentinel surveillance of COVID-19 should be based on the clinical specimens obtained through national sentinel surveillance of ILI/ARI/SARI.<br>• Representative stains of virus from different geographic locations and time points, as well as from patients of both genders, and across the age and severity spectrum should be selected for sequencing in order to monitor virus evolution and changes in the virus genome.<br>• Comprehensive surveillance. Countries comprehensively testing suspected cases for COVID-19 should monitor the number and proportion of hospitalized cases, cases admitted to intensive care units (ICU) or high-dependency units (HDU), and cases with fatal outcome among the number of confirmed cases.<br>• Hospital-based SARI surveillance. Countries no longer testing mild suspected cases for COVID-19 should at least test all severe acute respiratory infection (SARI) cases admitted to hospital and ICU/HDU, and monitor the proportion of confirmed COVID-19 cases among all SARI. This type of surveillance system can be comprehensive or based on a number of representative sentinel hospitals or areas. Sentinel hospitals should be selected if their catchment population is known and stable. A suitable alternative is to select all hospitals in a given area/region and use the population of that area/region as denominator.<br>• Mortality surveillance.<br>• Surveillance of hospitalized cases of COVID-19 with a focus on patient age, gender, medication, underlying<br>conditions, smoking and healthcare worker status, as well as ICU/HDU admission and clinical outcome. Regular descriptive and multivariable analysis should inform targeted preventative measures and messages. |
| Date not reported |  |  |  |  |
| New Zealand | Ministry of Health<sup>78</sup> | Scope: National<br>Setting: Community<br>Level of Evidence: Unclear | Wastewater surveillance<br>**Stated reason for use:** Wastewater testing is being used as an extra tool to help monitor for COVID-19. Wastewater testing may be able to give us an early warning of COVID-19 cases in the community. This will help to alert local communities to be more vigilant, keep up hygiene measures, and get tested and stay home if they are unwell. | Wastewater is collected from sites that contain a mixture of the wastewater from the toilets, sinks and drains of hundreds of thousands of people in a community. Wastewater is being sampled at least once a week from many sites around the country and the Institute of Environmental Research and Science (ESR) tests these samples at their lab to determine if they can detect the virus. |

